# Identification and Interaction Analysis of Molecular Markers in Pancreatic Ductal Adenocarcinoma by Integrated Bioinformatics Analysis and Molecular Docking Experiments

**DOI:** 10.1101/2020.12.20.20248601

**Authors:** Basavaraj Vastrad, Chanabasayya Vastrad, Anandkumar Tengli

## Abstract

The current investigation aimed to mine therapeutic molecular targets that play an key part in the advancement of pancreatic ductal adenocarcinoma (PDAC). The expression profiling by high throughput sequencing dataset profile GSE133684 dataset was downloaded from the Gene Expression Omnibus (GEO) database. Limma package of R was used to identify differentially expressed genes (DEGs). Functional enrichment analysis of DEGs were performed. Protein-protein interaction (PPI) networks of the DEGs were constructed. An integrated gene regulatory network was built including DEGs, microRNAs (miRNAs), and transcription factors. Furthermore, consistent hub genes were further validated. Molecular docking experiment was conducted. A total of 463 DEGs (232 up regulated and 231 down regulated genes) were identified between very early PDAC and normal control samples. The results of Functional enrichment analysis revealed that the DEGs were significantly enriched in vesicle organization, secretory vesicle, protein dimerization activity, lymphocyte activation, cell surface, transferase activity, transferring phosphorus-containing groups, hemostasis and adaptive immune system. The PPI network and gene regulatory network of up regulated genes and down regulated genes were established, and hub genes were identified. The expression of hub genes (CCNB1, FHL2, HLA-DPA1 and TUBB1) were also validated to be differentially expressed among PDAC and normal control samples. Molecular docking experiment predicted the novel inhibitory molecules for CCNB1 and FHL2. The identification of hub genes in PDAC enhances our understanding of the molecular mechanisms underlying the progression of this disease. These genes may be potential diagnostic biomarkers and/or therapeutic molecular targets in patients with PDAC.

## Introduction

Pancreatic ductal adenocarcinoma (PDAC) is one of the most prevalent cancers in the world and primary tumor of the pancreas [1]. PDAC is a global burden ranking 15th in terms of incidence and fourth in terms of mortality [2]. Despite new developments in multimodal therapy its overall 5-year survival rate remains less than 8% [3]. PDAC treatment commonly includes surgery, radiation, chemotherapy and immunotherapy [4]. However, PDAC remains common and malignant due to recurrence and metastasis, and ultimately key cause of PDAC associated death [5]. Therefore, there is a vital need to advance new diagnostic strategies and therapeutic agents to upgrade the prognosis of patients with PDAC.

The molecular mechanisms of PDAC tumorigenesis and development remain imprecise. It is therefore key to identify novel genes and pathways that are linked with PDAC tumorigenesis and patient prognosis, which may not only help to illuminate the underlying molecular mechanisms involved, but also to disclose novel diagnostic markers and therapeutic targets. Oji et al [6] demonstrated that the over expression of WT1 is linked with prognosis in patients with PDAC. A previous investigation reported that phosphoinositide 3-kinase signaling pathway is linked with development of PDAC [7]. Expression profiling by high throughput sequencing can promptly uncover gene expression on a global basis and are specially useful in identifying for differentially expressed genes (DEGs) [8]. A huge amount of data has been generated through the use of microarrays and the majority of such data has been deposited and saved in public databases. Previous investigations concerning PDAC gene expression profiling have diagnosed hundreds of DEGs [9].

The aim of this investigation was to identify hub genes and pathways in PDAC using bioinformatics methods. Our investigation contributes predictable biomarkers for early detection and prognosis, as well as effective drug targets for treating PDAC.

## Materials and methods

### Sequencing data

PDAC expression profiling by high throughput sequencing dataset in this investigation was downloaded from the GEO database (https://www.ncbi.nlm.nih.gov/gds/) [10]. The DEGs were considered by 1 independent PDAC dataset, GSE133684 [11] with 284 PDAC and 117 normal samples. The GSE133684 expression profiling by high throughput sequencing data was based on the GPL20795 HiSeq X Ten (Homo sapiens) platform..

### Identification of DEGs

The Limma package of R language were used to normalize and convert the raw data to expression profiles [12]. The limma package of R language was used for DEGs between PDAC and normal control samples [12]. The P-value was adjusted by the Benjamini-Hochberg method [13]. An adjusted P-value <0.05 and |log2 fold change (FC) | >1 were considered as threshold values for DEGs identification. The ggplot2 package and gplots package of R language was used to generate volcano plot and heat map. The identified DEGs were preserved for further bioinformatics analysis.

### GO analysis and pathway enrichment analysis of DEGs

The GO repository (http://geneontology.org/) [14] consists of a massive set of annotation terms and is generally used for annotating genes and identifying the distinctive biological aspects for expression profiling by high throughput sequencing data. The REACTOME database (https://reactome.org/) [15] contains data on known genes and their biochemical functions and is used for identifying functional and metabolic pathways. By performing the GO and REACTOME enrichment analysis at the functional level, we can boost a better understanding of the roles of these DEGs in the induction and in the advancement of PDAC. The ToppGene (ToppFun) (https://toppgene.cchmc.org/enrichment.jsp) [16] is an online resource that add tools for functional annotation and bioinformatics analysis. Both GO categories and REACTOME pathway enrichment analysis were implemented using ToppGene to inform the functions of these DEGs. P<0.05 was considered to indicate a statistically significant difference.

### Protein-protein interaction (PPI) network construction and module analysis

The online database IID interactome (http://iid.ophid.utoronto.ca//)[17] was used to construct a PPI network of the proteins encoded by DEGs. Then, Cytoscape software (Version 3.8.1) [18] was applied to perform protein interaction association network analysis and analyze the interaction correlation of the candidate proteins encoded by the DEGs in PDAC. Next, the Network Analyzer Cytoscape plug-in was applied to calculate node degree [19], betweenness centrality [20], stress centrality [21], closeness centrality [22]. Finally, the PEWCC1 (http://apps.cytoscape.org/apps/PEWCC1) [23] module for Cytoscape was used to collect the significant modules in the PPI network complex.

### Construction of miRNA-DEG regulatory network

The miRNet database (https://www.mirnet.ca/) [24] is a database, containing miRNAs involved in various diseases. The miRNAs related to PDAC were searched from miRNet database. Through getting the intersection of the miRNAs and the DEGs, the miRNA-DEG regulatory relationships were selected. Finally, miRNA-DEG regulatory network was built using Cytoscape software.

### Construction of TF-DEG regulatory network

The NetworkAnalyst database (https://www.networkanalyst.ca/) [25] is a database, containing TFs involved in various diseases. The TFs related to PDAC were searched from TF database. Through getting the intersection of the TFs s and the DEGs, the TF-DEG regulatory relationships were selected. Finally, TFs-DEG regulatory network was built using Cytoscape software.

### Hub genes validation

After hub genes identified from expression profiling by high throughput sequencing dataset, UALCAN (http://ualcan.path.uab.edu/analysis.html) [26] was used to validate the selected up regulated and down regulated hub genes. UALCAN is an online tool for gene expression analysis between PDAC and normal data from The Cancer Genome Atlas (TCGA). It adds data such as gene expression, tumor staging, and survival period for PDAC. cBioPortal is an online platform (http://www.cbioportal.org) [27] for gene alteration of hub genes analysis from TCGA. Human protein atlas is an online database (HPA, www.proteinatlas.org) [28] for protein expression analysis between PDAC and normal data from TCGA. TIMER is an online platform (https://cistrome.shinyapps.io/timer/) [29] for immune infiltration analysis from TCGA. To explore diagnostic biomarkers of PDAC, we used the above hub genes as candidates to find their diagnostic value based on generalized linear model (GLM) [30]. The pROC in R was used for Receiver operating characteristic (ROC) curve analysis [30]. In brief, half of the samples (PDAC = 142, controls = 59) were aimlessly distributed as the training set and remaining data were used as the test set, which were used to set up a model. An ROC curve analysis was tested to calculate the specificity and sensitivity of the GLM prediction model. The area under the curve (AUC) was figure out to determine the diagnostic efficiency of the classifier.

### RT-PCR analysis

TRI Reagent® (Sigma, USA) was used to extract total RNA from the culture cells of PDAC (CRL-2549™) and normal (CRL-2989™) according to the manufacturer’s protocol. Reverse transcription cDNA kit (Thermo Fisher Scientific, Waltham, MA, USA) and random primers were used to synthesize cDNA. Quantitative real-time PCR (qRT-PCR) was conducted on the 7 Flex real-time PCR system (Thermo Fisher Scientific, Waltham, MA, USA). The reaction guideline included a denaturation program (5 min at 95 °C), followed by an amplification and quantification program over 40 cycles (15 s at 95°C and 45 s at 65°C). Each sample was tested in triplicates. Table 1 depicts the primer sequences of hub genes. The expression level was resolved as a ratio between the hub genes and the internal control β-actin in the same mRNA sample, and determined by the comparative CT method [31]. Levels of CCNB1, FHL2, HLA-DPA1 and TUBB1 expression were determined by the 2^−^ΔΔCt method.

**Table 1.**
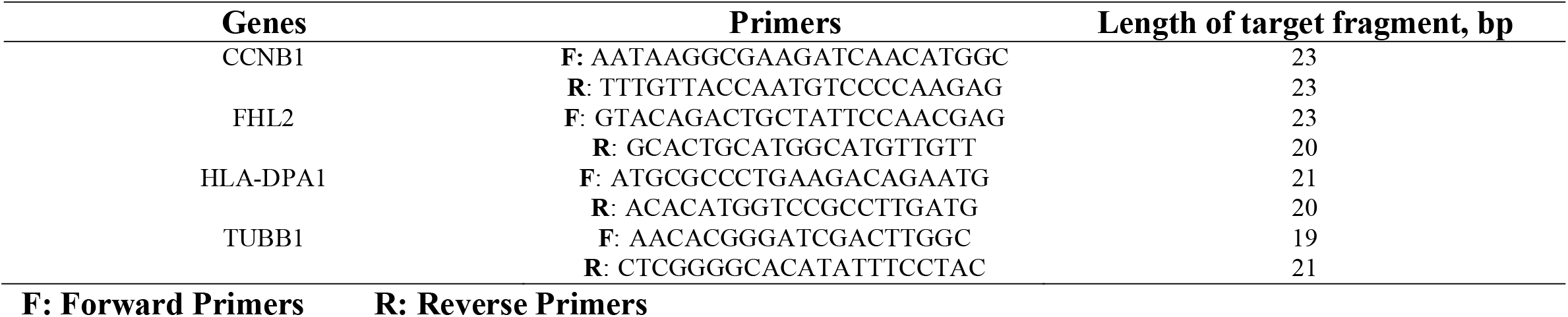
The sequences of primers for quantitative RT-PCR

### Molecular docking experiment

The module SYBYL-X 2.0 perpetual software were used for Surflex-Docking of the designed molecules. The molecules were sketched by using ChemDraw Software and imported and saved in sdf. format using openbabel free software. The protein structures of CyclinB1 (CCNB1) its co-crystallisedprotein of PDB code 4Y72, 5H0V and Four and half LIM domains 2 (FHL2) its NMR structure of proteins 2D8Z and 2EHE was retrieved from Protein Data Bank [32-34]. Together with the TRIPOS force field, GasteigerHuckel (GH) charges were added to all designed derivatives for the structure optimization process.In addition, energy minimization was carried outusing MMFF94s and MMFF94 algorithm process. Protein processing was carried out after the incorporation of protein.The co-crystallized ligand and all water molecules were removed from the crystal structure; more hydrogens were added and the side chain was set. TRIPOS force field was used for the minimization of structure. The compounds’ interaction efficiency with the receptor was represented by the Surflex-Dock score in kcal / mol units. The interaction between the protein and the ligand, the best pose was incorporated into the molecular area. The visualisation of ligand interaction with receptor is done by using discovery studio visualizer.

## Results

### Identification of DEGs

We analyzed the DEGs of GSE133684 by using the limma package. We used p□<□0.05 and |logFC|□≥□1 as the cutoff criteria. We screened 463 DEGs, including 232 up regulated genes and 231 down regulated genes in PDAC samples compared with normal control samples and are listed in Table 2. We identified all the DEGs which were shown in the above volcano map according to the value of |logFC| is shown in Fig. 1 and then displayed the DEGs on a heatmap is shown in Fig. 2.

**Table 2.**
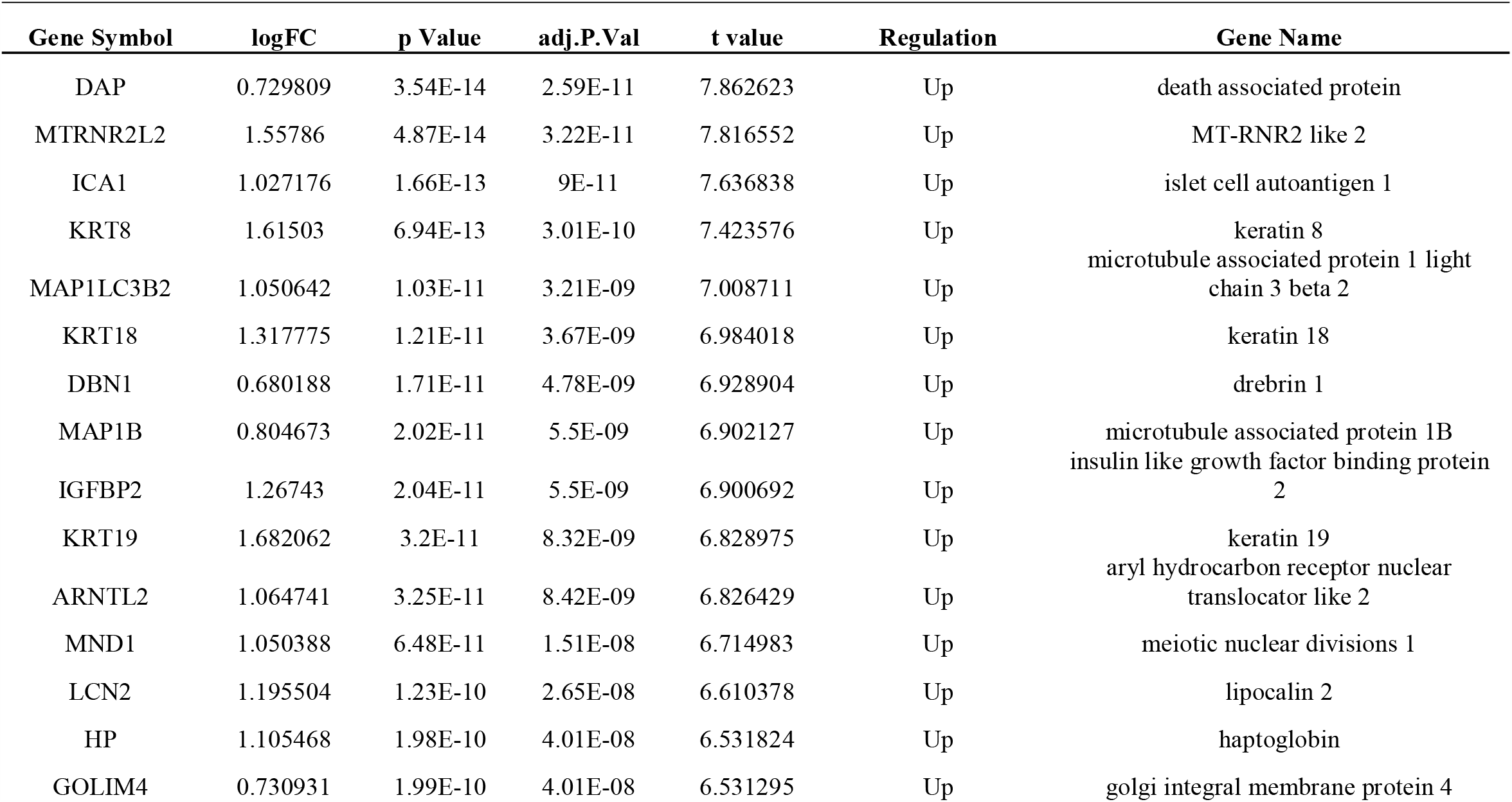
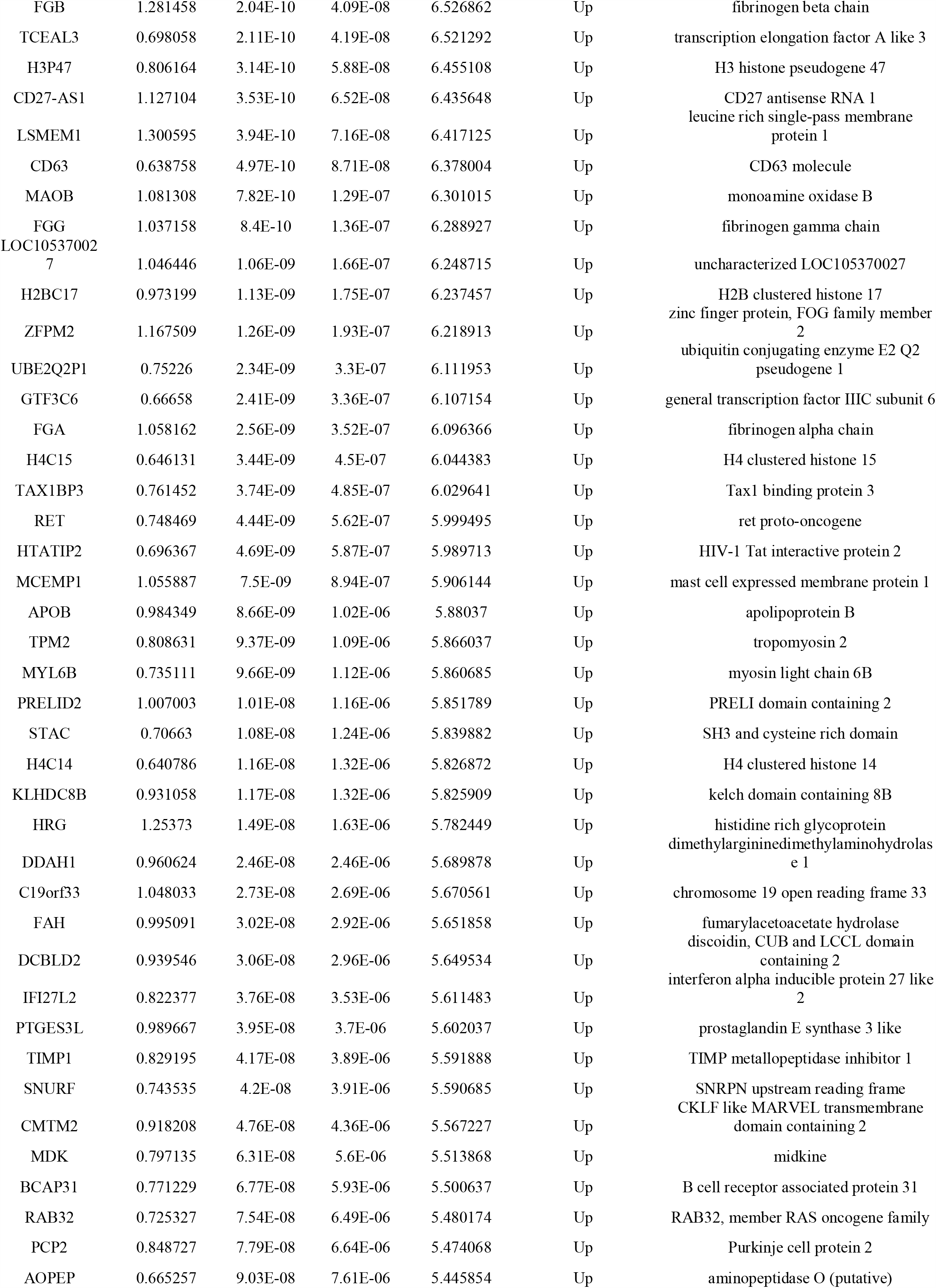
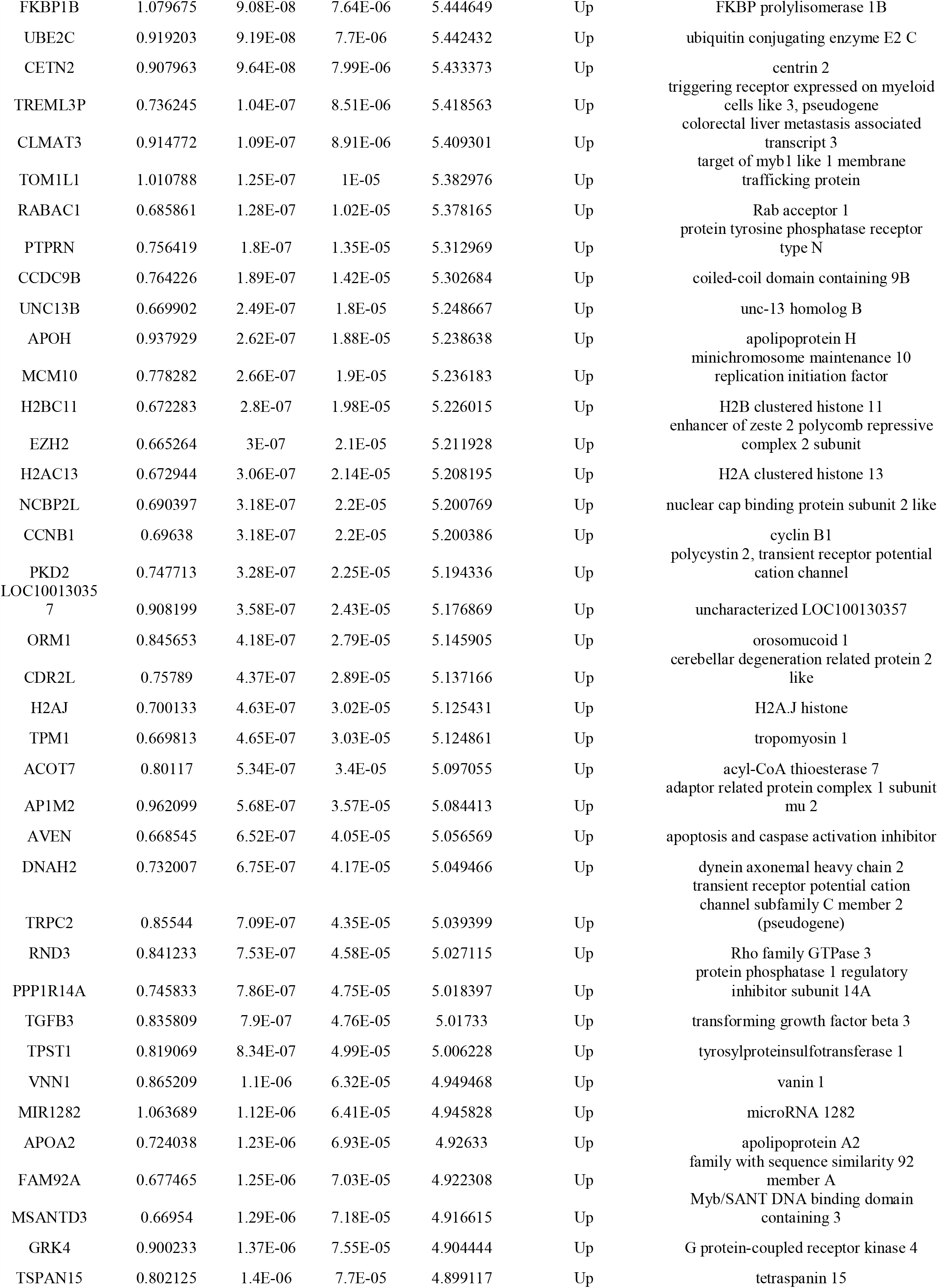
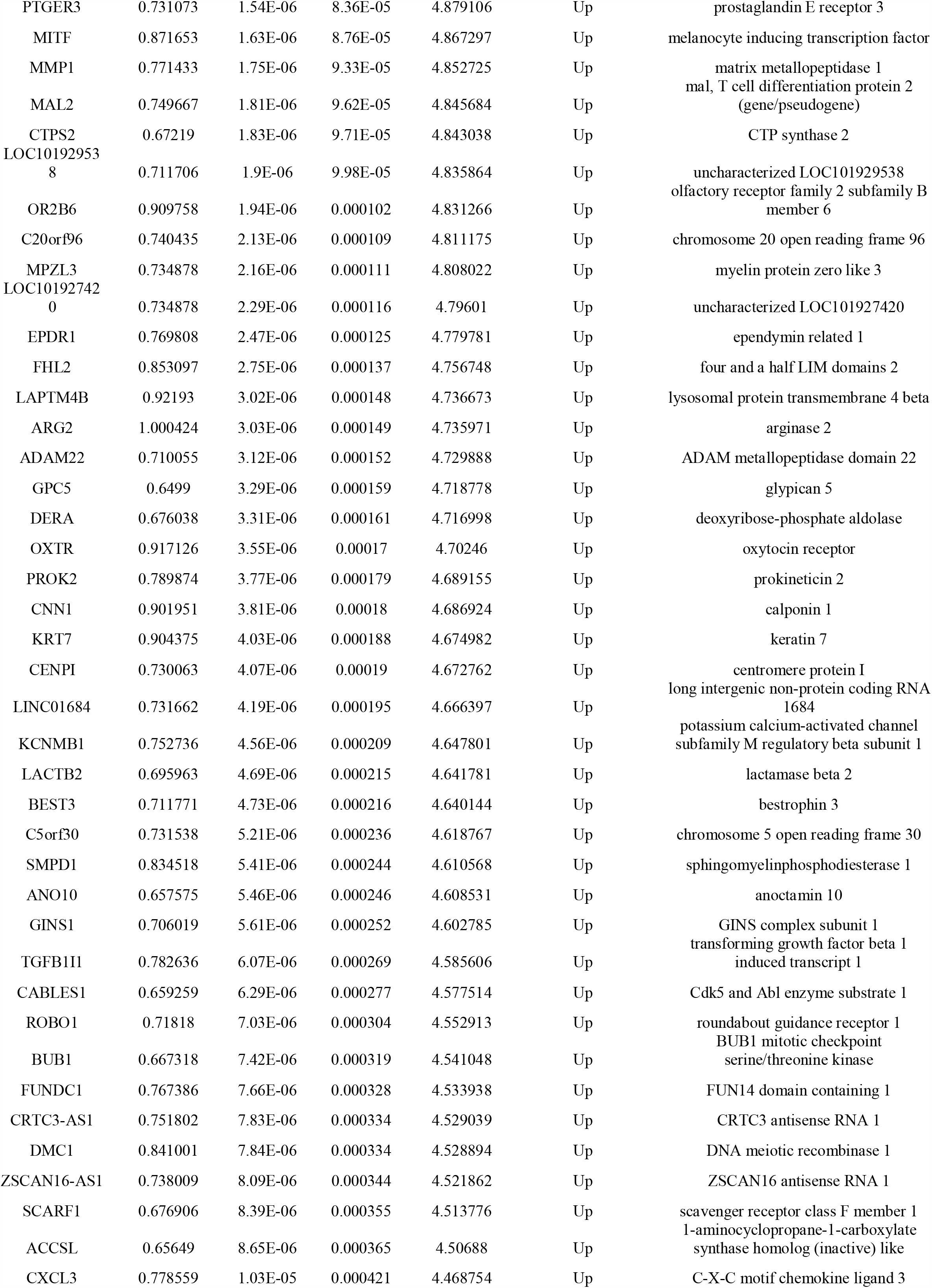
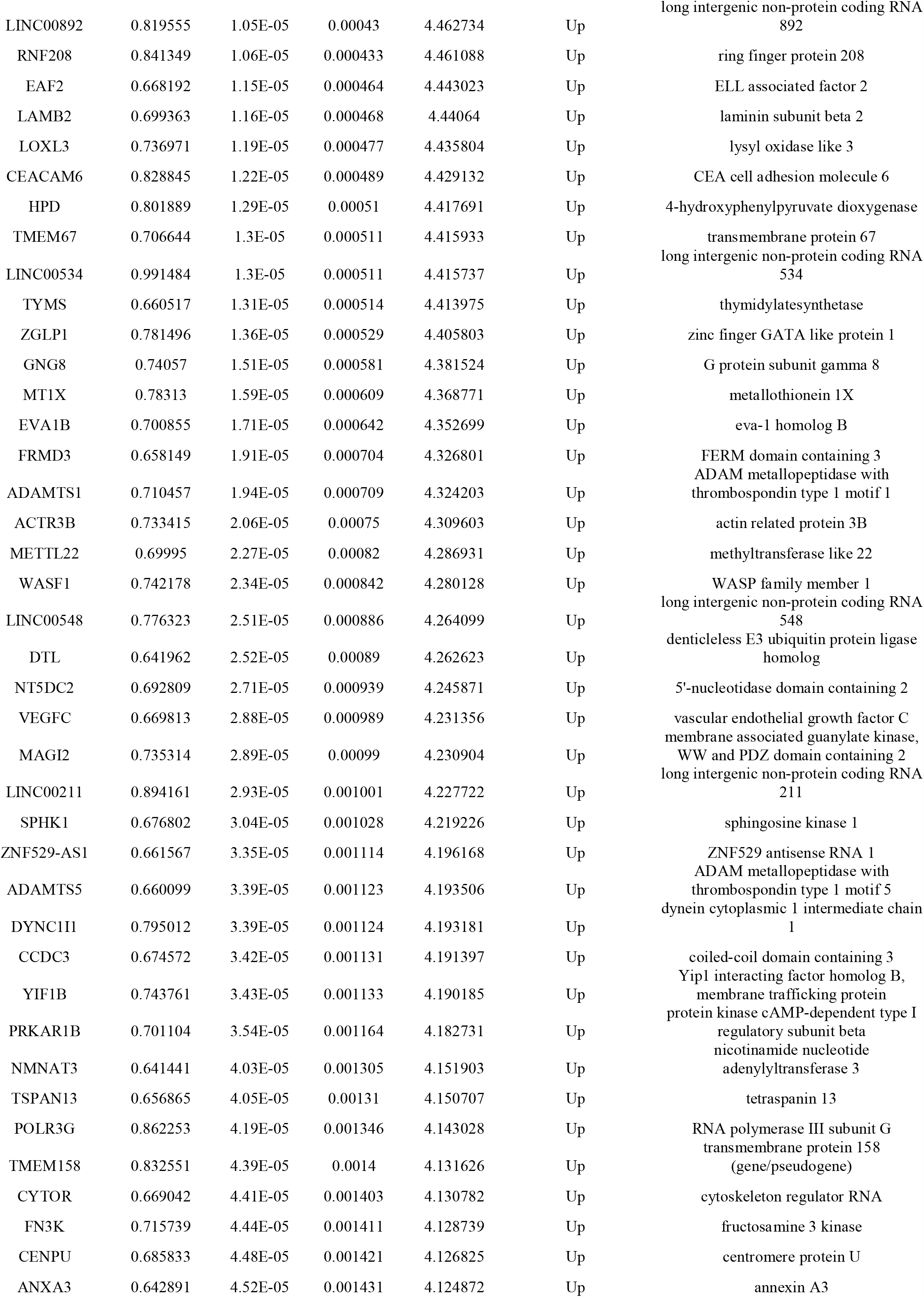
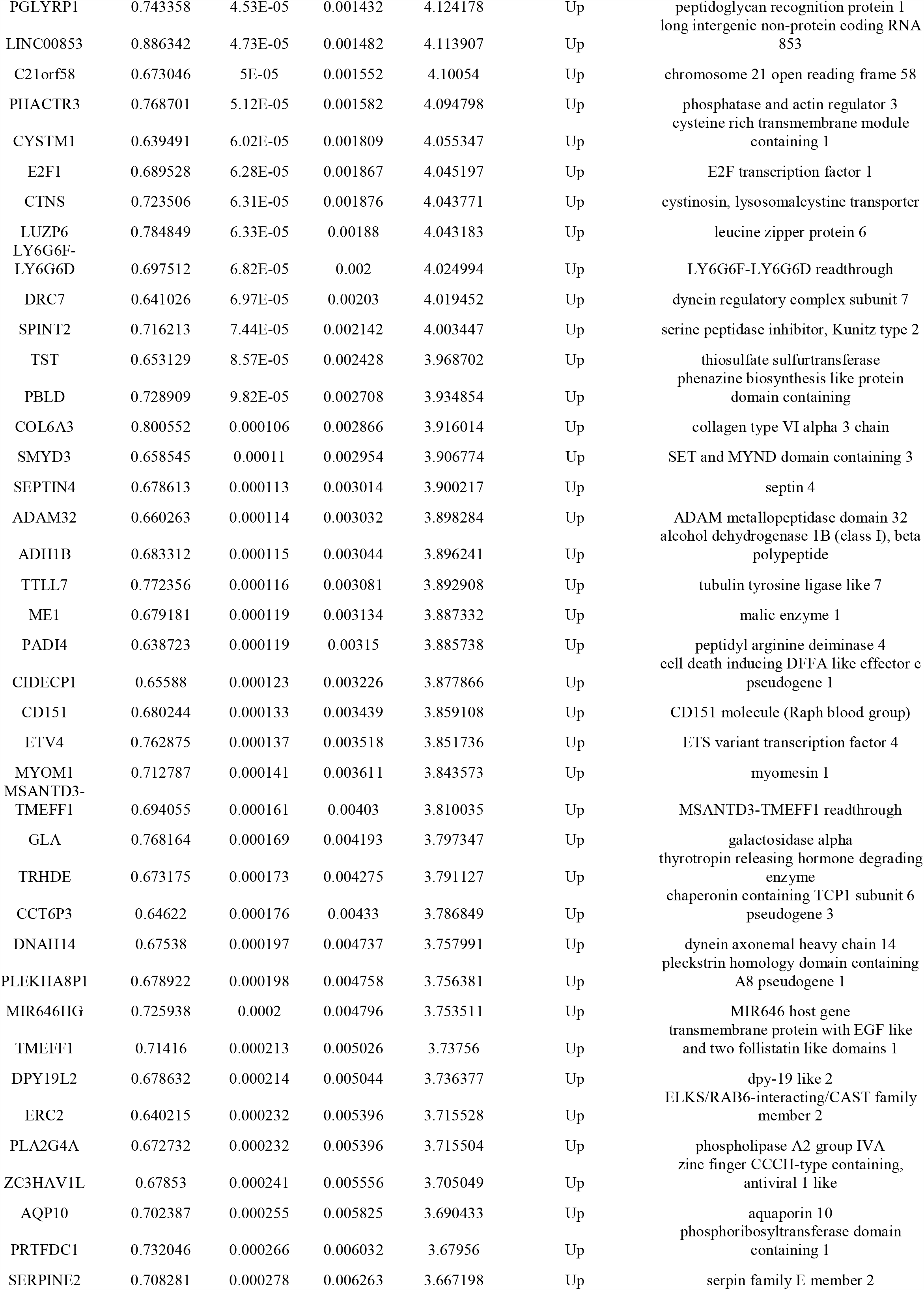
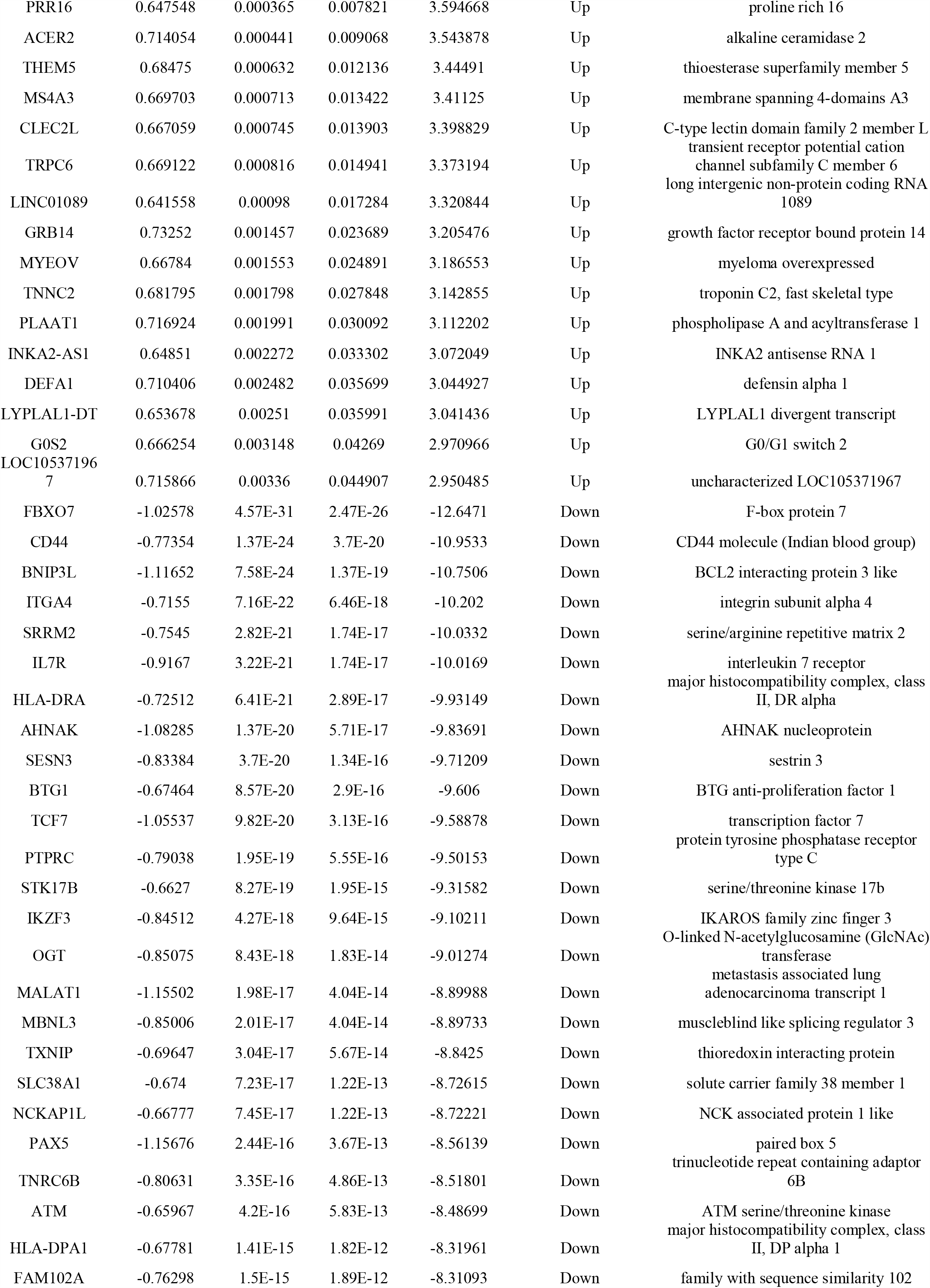
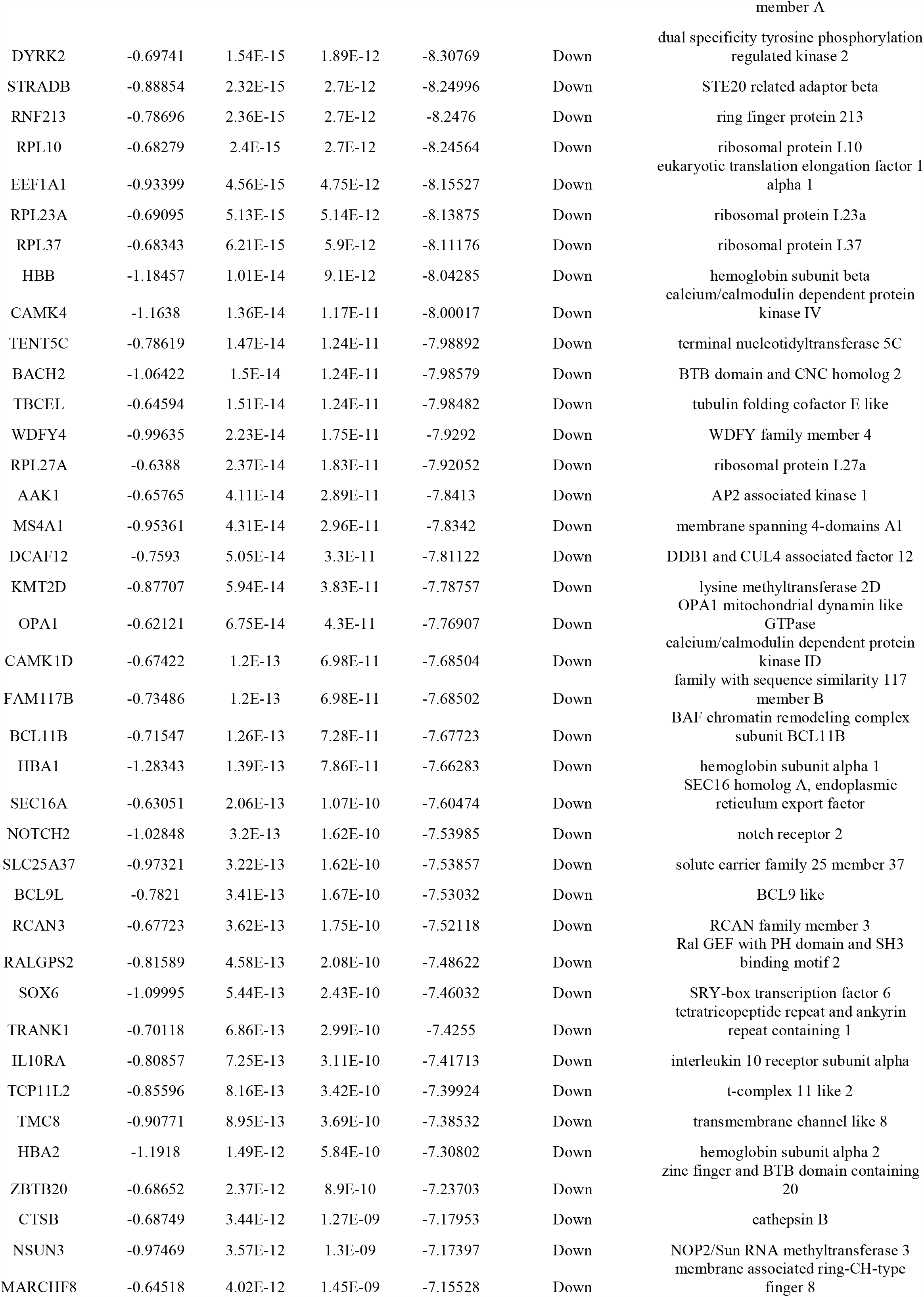
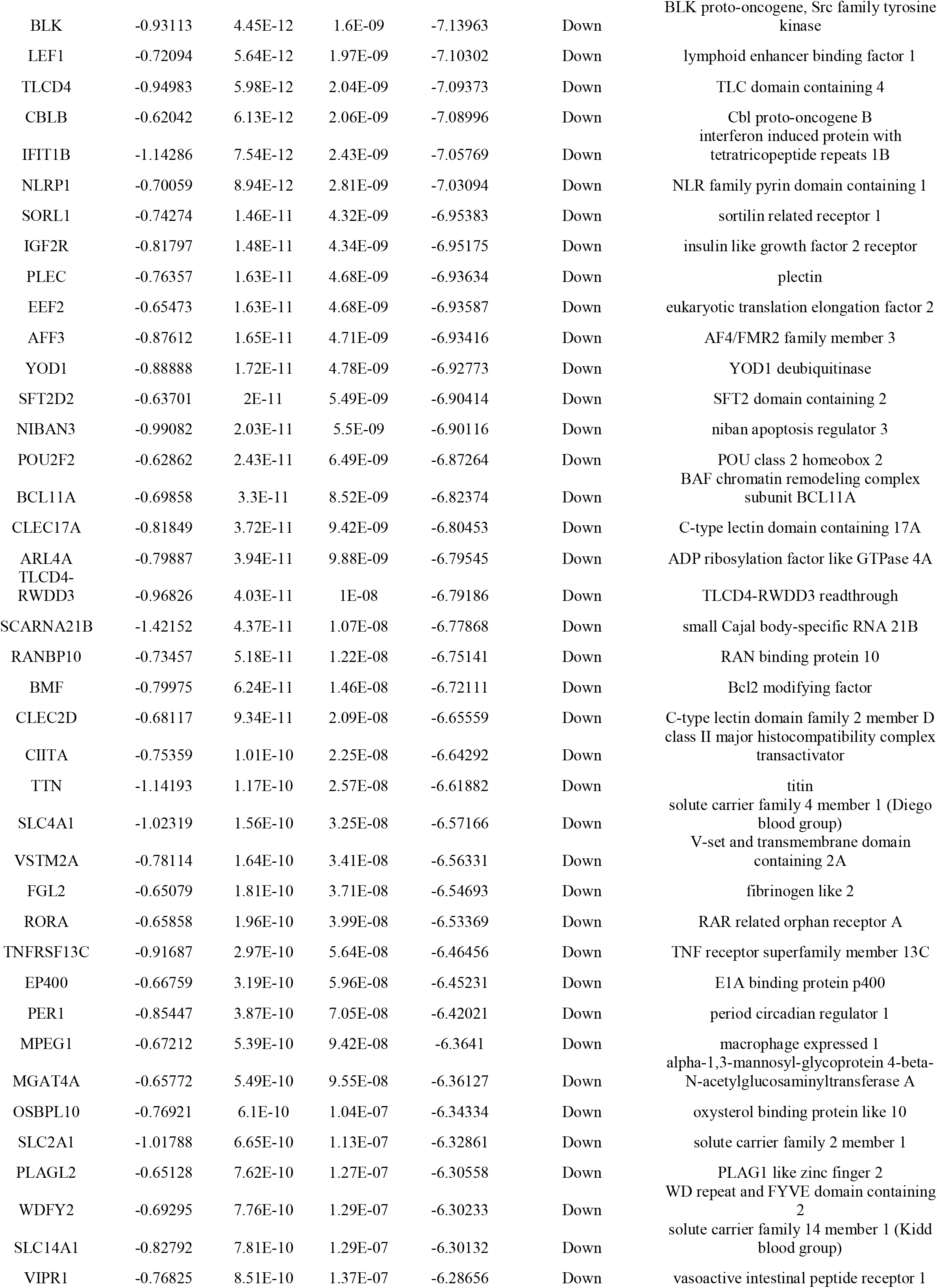
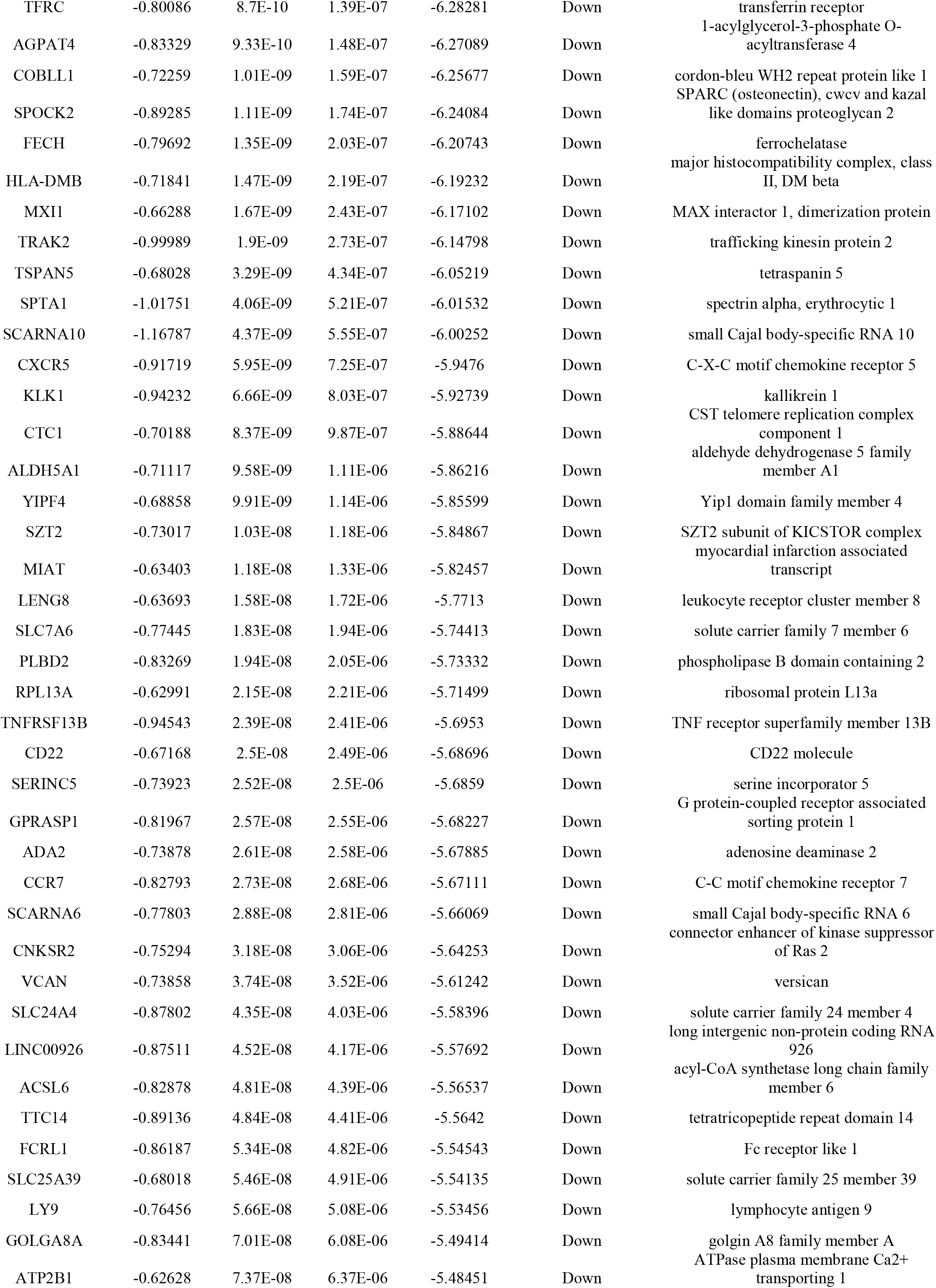
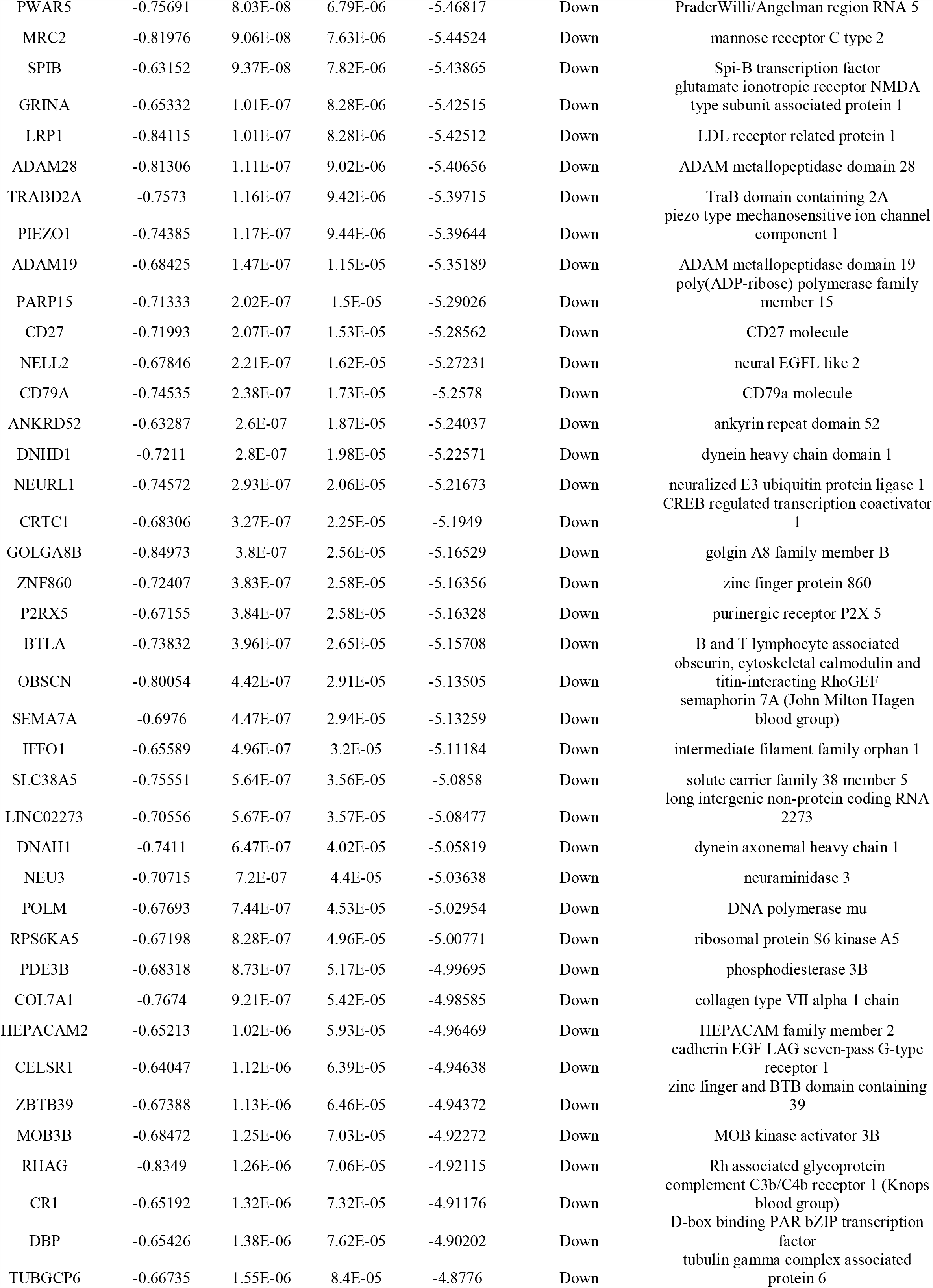
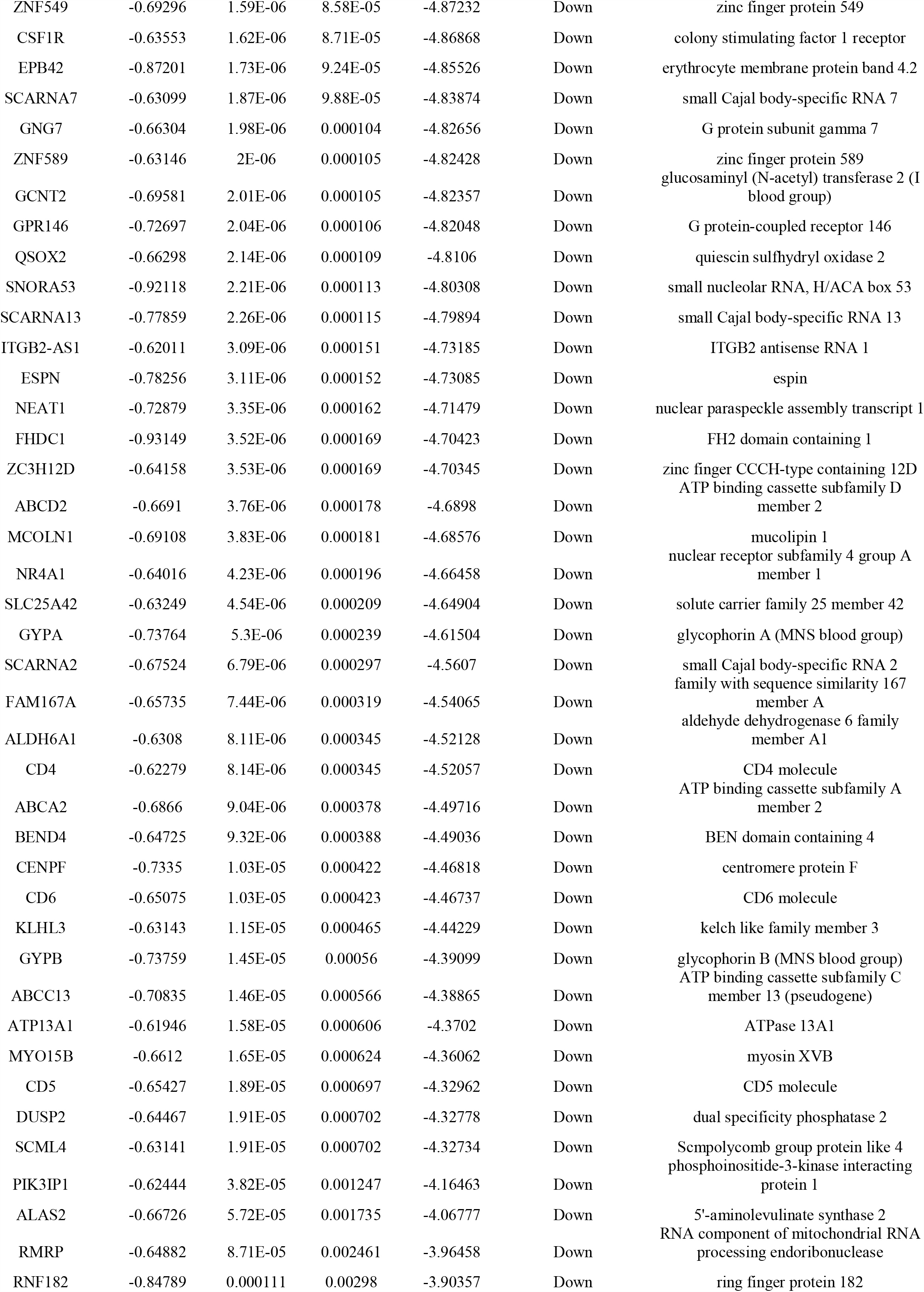
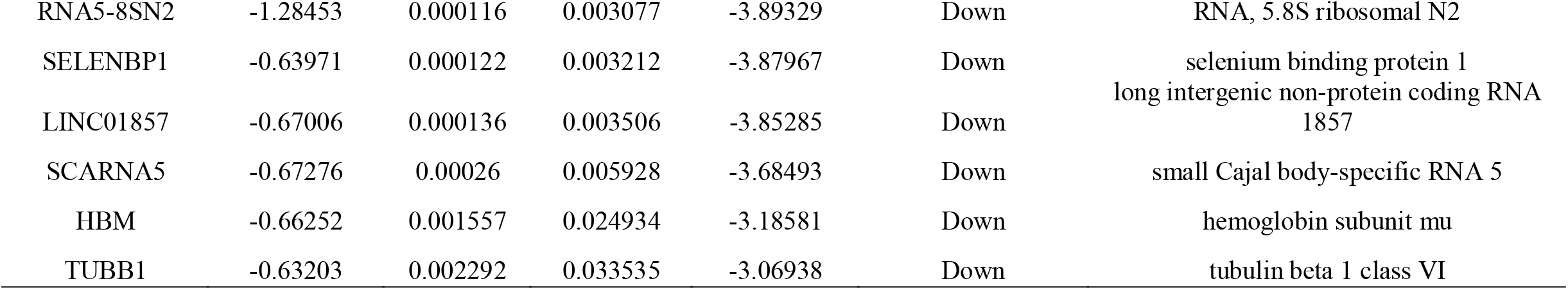
The statistical metrics for key differentially expressed genes (DEGs)

**Fig. 1.**
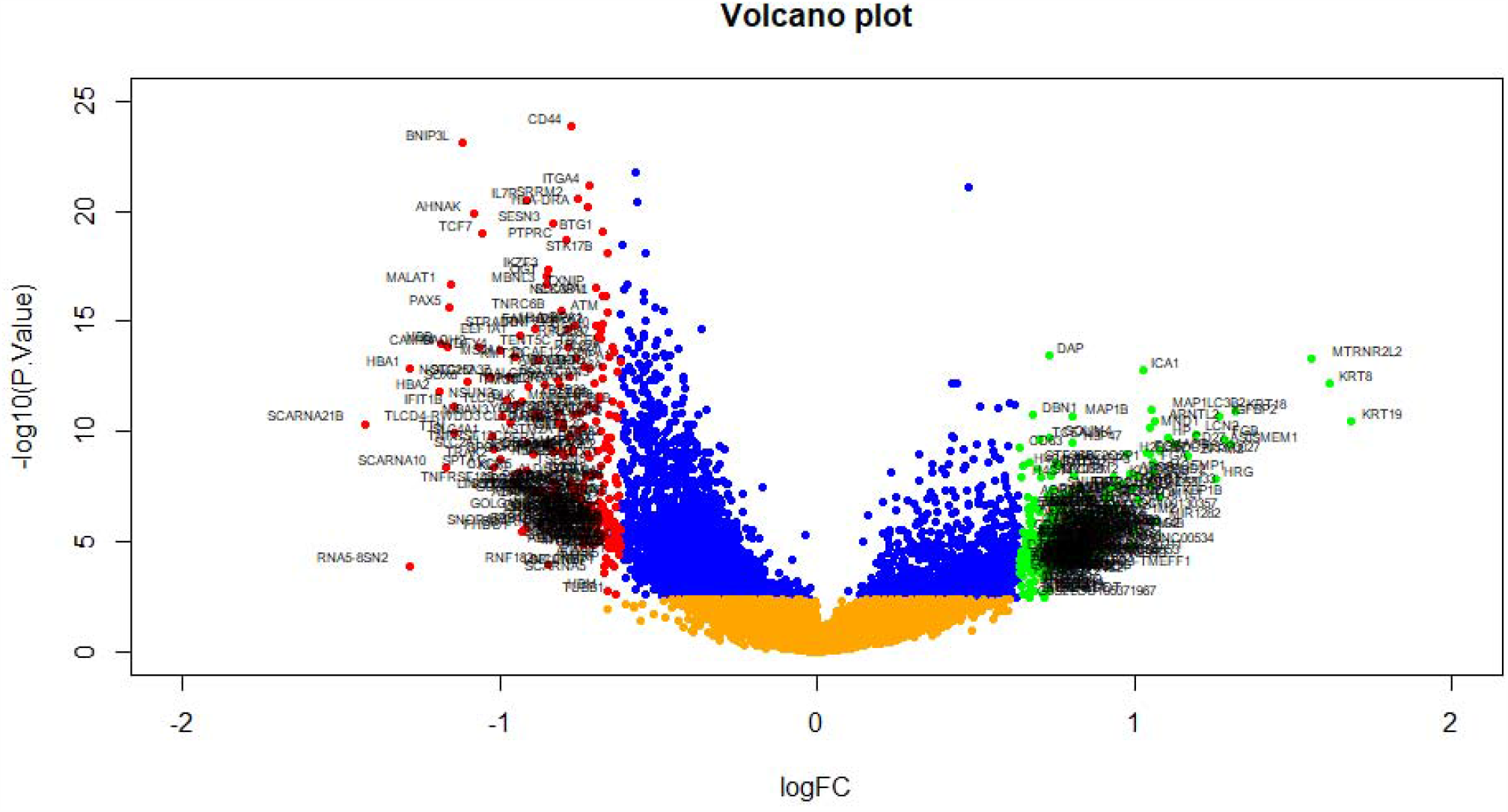
Volcano plot of differentially expressed genes. Genes with a significant change of more than two-fold were selected. Green dot represented up regulated significant genes and red dot represented down regulated significant genes.

**Fig. 2.**
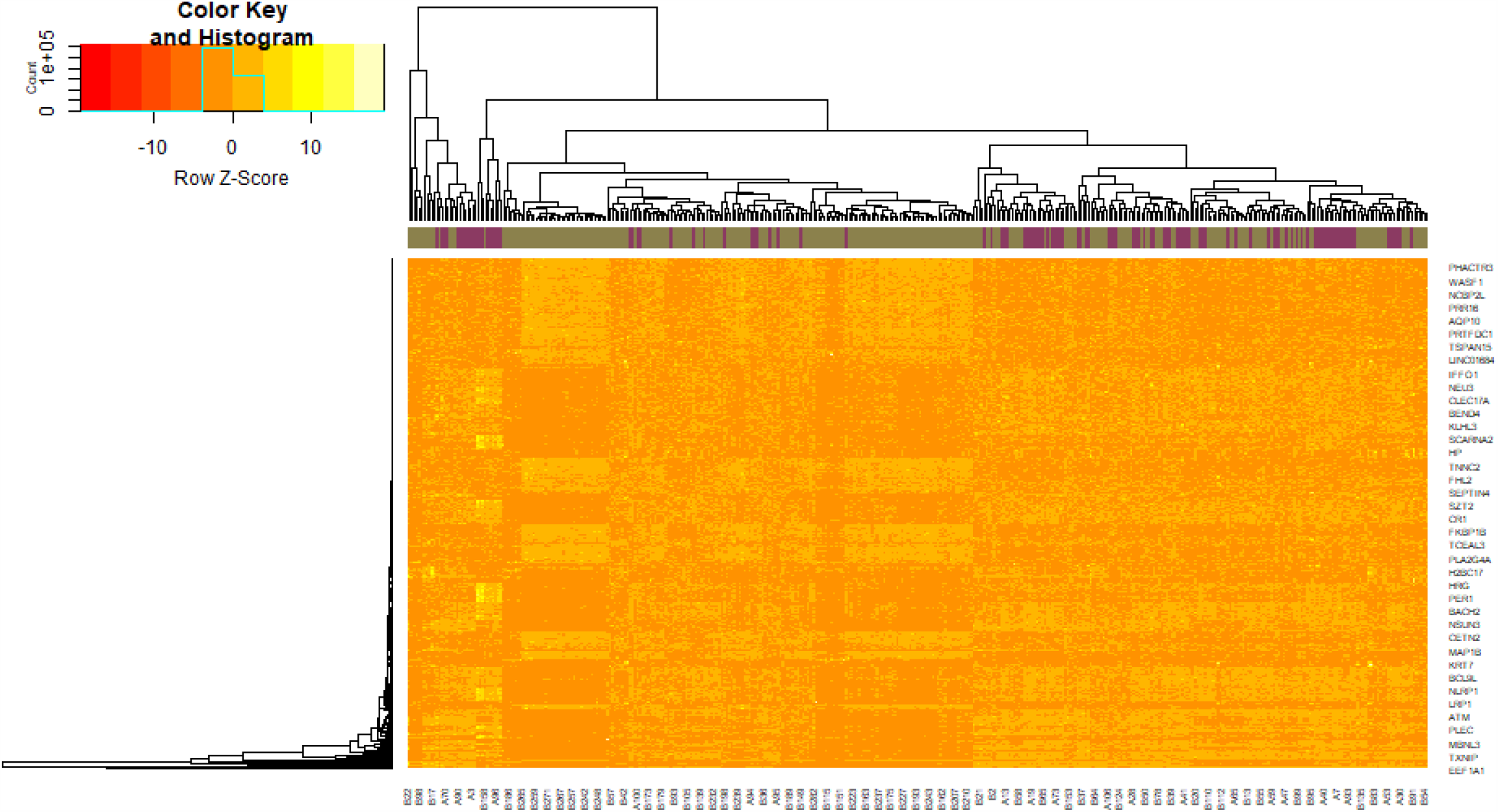
Heat map of differentially expressed genes. Legend on the top left indicate log fold change of genes. (A1 – A114 = normal control samples; B1 – B284 = PDAC samples)

### GO analysis and pathway enrichment analysis of DEGs

To symbolize the function of the DEGs and to identify important candidate pathways, GO functional enrichment analysis and REACTOME pathway enrichment analysis were performed. The results of GO categories analysis including biological processes (BP), cellular components (CC) and molecular functions (MF) are listed in Table 3. Firstly, the up regulated genes were annotated with the BP category, including vesicle organization and secretion, whereas the down regulated genes were annotated with the GO terms, including lymphocyte activation and regulation of cell death. Secondly, the up regulated genes were annotated with the GO terms of the CC category, namely secretory vesicle and whole membrane, whereas the down regulated genes were annotated with the GO terms, including cell surface and intrinsic component of plasma membrane.

**Table 3.**
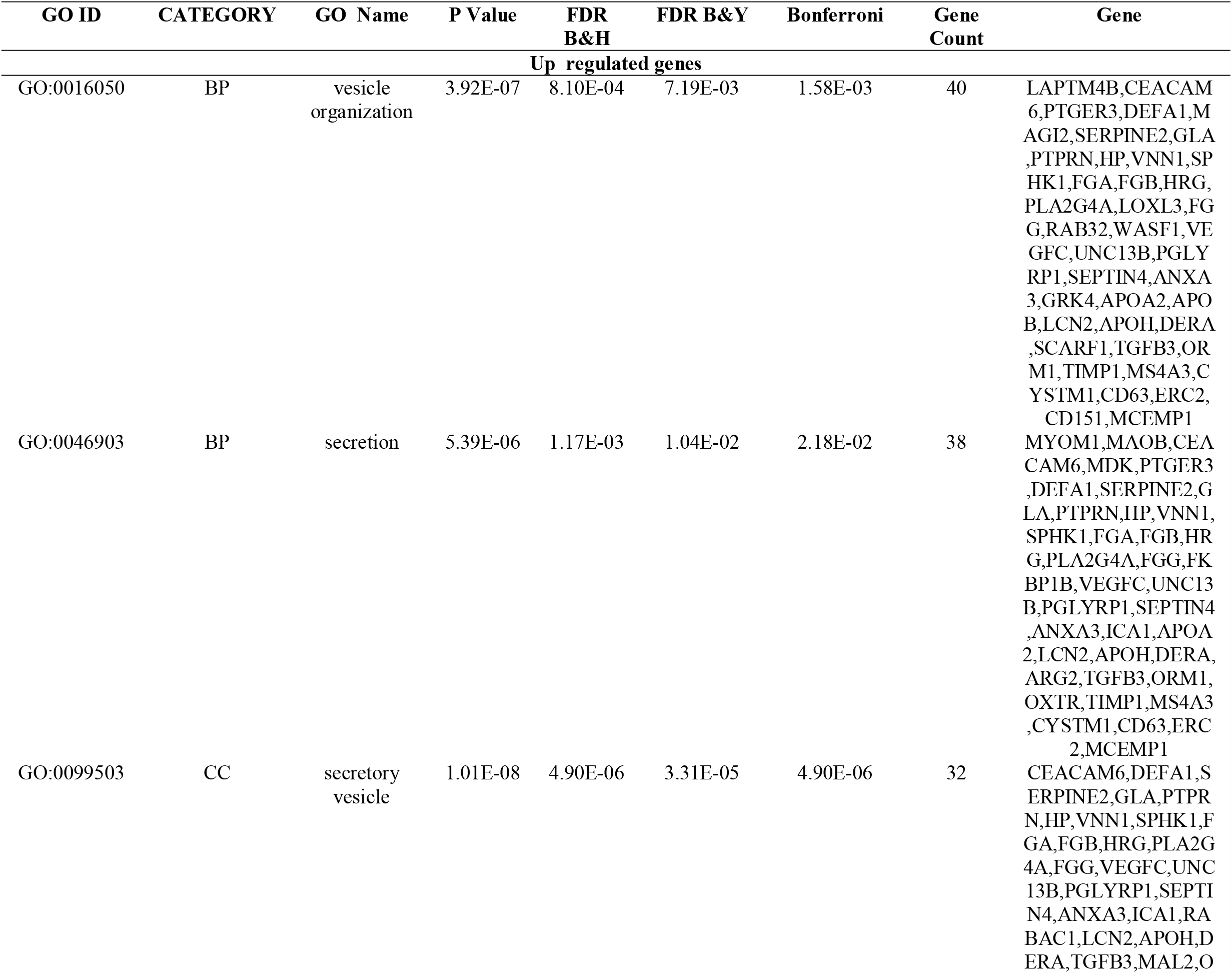
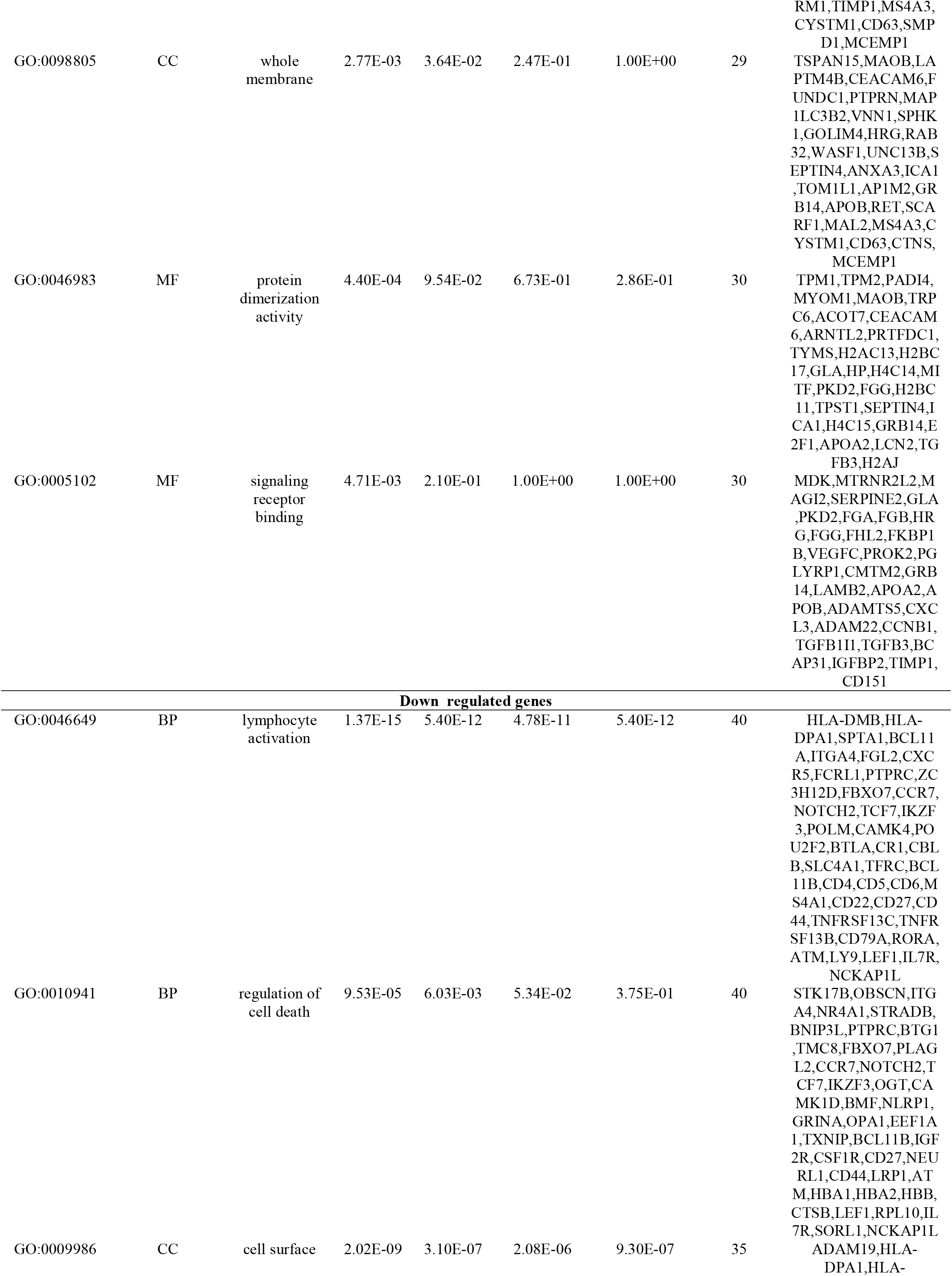
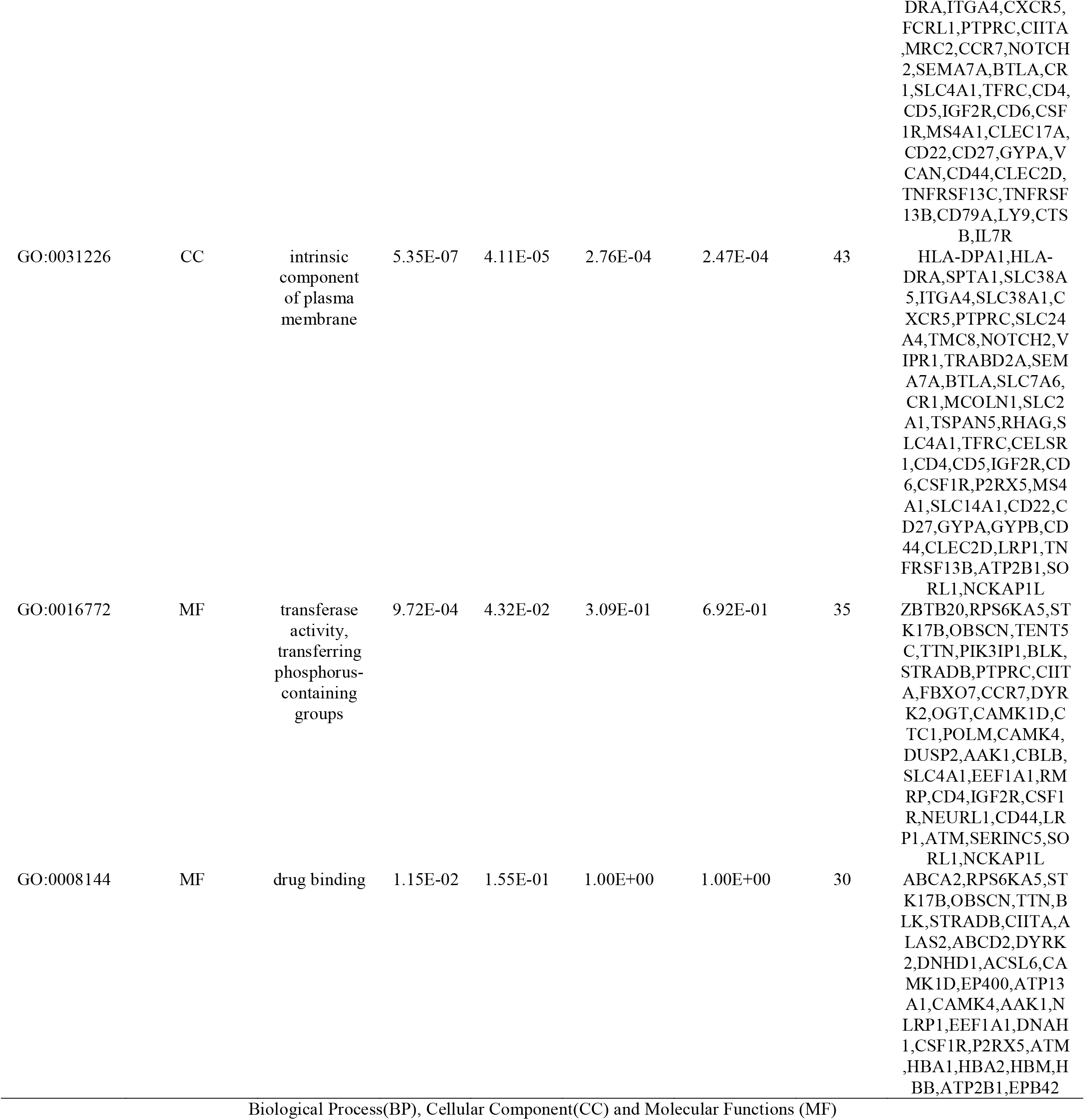
The enriched GO terms of the up and down regulated differentially expressed genes

Thirdly, the up regulated genes were annotated with the GO terms of the MF category, such as protein dimerization activity and signaling receptor binding, whereas the down regulated genes were annotated with the GO terms, including transferase activity, transferring phosphorus-containing groups and drug binding. As shown in Table 4, the significantly enriched REACTOME pathways of the up regulated genes with P<0.05 were hemostasis and cell cycle, whereas down regulated genes with P<0.05 were adaptive immune system and transmembrane transport of small molecules.

**Table 4.**
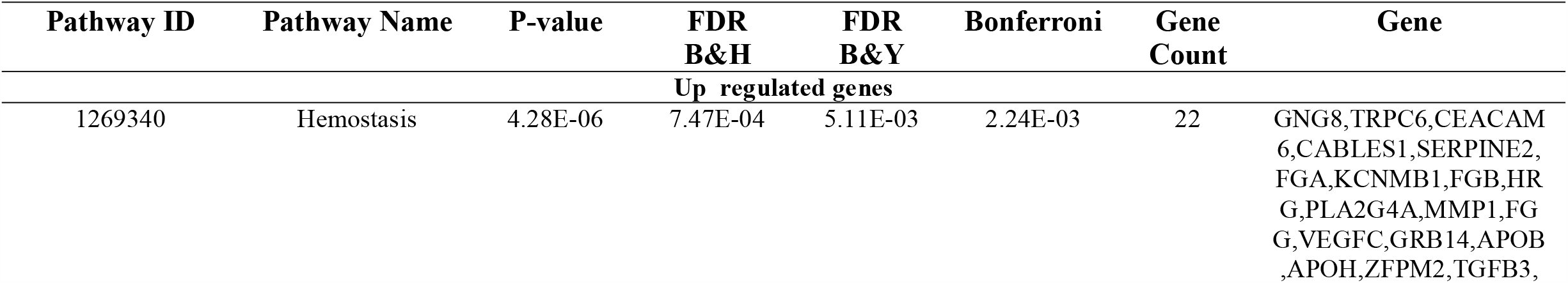
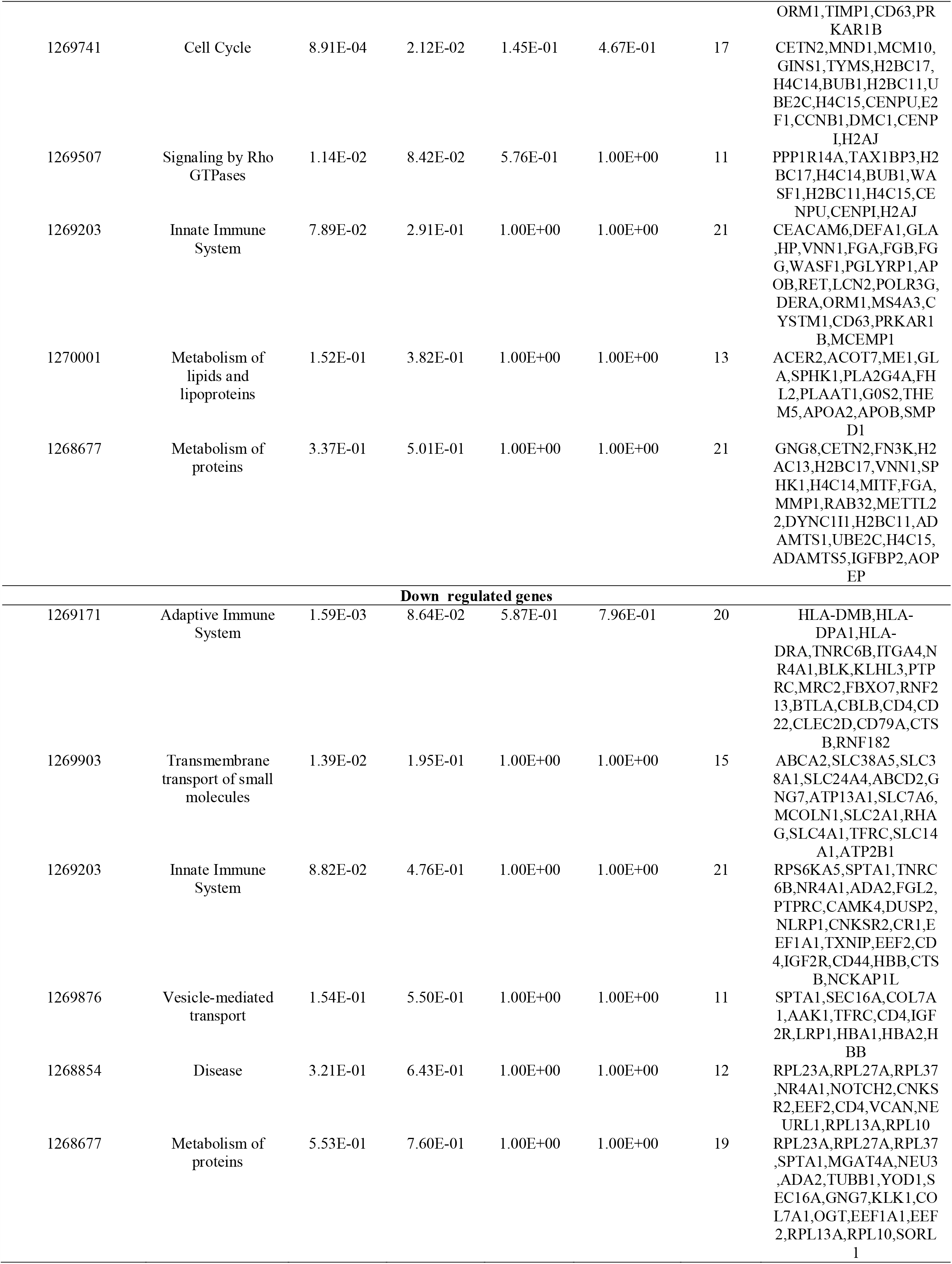
The enriched pathway terms of the up and down regulated differentially expressed genes

### Protein-protein interaction (PPI) network construction and module analysis

After all the DEGs were uploaded to the online IID interactome database, the PPI network with 6188 nodes and 13153 edges was constructed using the Cytoscape software (Fig. 3A). Hub DEGs with the node degree, betweenness centrality, stress centrality and closeness centrality were obtained and are listed in Table 5. Among them, CCNB1 and FHL2 were the major up regulated genes, while HLA-DPA1 and TUBB1 were the major down regulated genes. Then, two significant module that fulfilled the cut-off criteria, namely, PEWCC1 scores >3 and number of nodes >5, was screened (Fig. 3B and Fig. 3C). The FGB, FGA, FGG, EEF1A1, RPL13A, ITGA4, RPL27A, RPL23A and RPL10 genes were identified in these modules. GO analysis of these genes showed that they were annotated with vesicle organization, regulation of cell death and lymphocyte activation. In addition, the REACTOME enrichment analysis suggested that these genes were mainly involved in hemostasis, innate immune system, disease and adaptive immune system.

**Table 5.** Topology table for up and down regulated genes.

| Regulation | Node | Degree | Betweenness | Stress | Closeness |
| --- | --- | --- | --- | --- | --- |
| Up | EZH2 | 350 | 0.076224 | 81130962 | 0.349924 |
| Up | DBN1 | 256 | 0.049032 | 39941040 | 0.348151 |
| Up | CCNB1 | 158 | 0.024471 | 43653864 | 0.313488 |
| Up | FHL2 | 158 | 0.031673 | 16397718 | 0.336762 |
| Up | E2F1 | 151 | 0.025971 | 16601668 | 0.333208 |
| Up | TPM1 | 134 | 0.022043 | 16976362 | 0.329604 |
| Up | KRT18 | 132 | 0.018793 | 17362522 | 0.338383 |
| Up | TPM2 | 126 | 0.021531 | 17717608 | 0.323503 |
| Up | FGB | 115 | 0.021143 | 15804232 | 0.298889 |
| Up | PTGER3 | 113 | 0.019531 | 25585198 | 0.284342 |
| Up | SPINT2 | 108 | 0.023415 | 14310762 | 0.303448 |
| Up | BUB1 | 102 | 0.018982 | 9083534 | 0.319527 |
| Up | PTPRN | 93 | 0.015139 | 21217898 | 0.283912 |
| Up | BCAP31 | 91 | 0.017733 | 11925460 | 0.323825 |
| Up | MAP1B | 91 | 0.011535 | 22589676 | 0.310858 |
| Up | KRT8 | 79 | 0.009251 | 9231600 | 0.332081 |
| Up | TMEM67 | 76 | 0.014065 | 4982016 | 0.290838 |
| Up | RABAC1 | 74 | 0.013172 | 9708740 | 0.293098 |
| Up | APOB | 72 | 0.011818 | 15182272 | 0.302587 |
| Up | UBE2C | 69 | 0.00877 | 8892764 | 0.298716 |
| Up | SPHK1 | 68 | 0.011494 | 4951244 | 0.325666 |
| Up | TOM1L1 | 66 | 0.00895 | 6727694 | 0.305124 |
| Up | RET | 66 | 0.009723 | 10964184 | 0.305199 |
| Up | WASF1 | 65 | 0.010329 | 14175172 | 0.29873 |
| Up | KRT19 | 64 | 0.008418 | 5490710 | 0.32224 |
| Up | CENPU | 61 | 0.011081 | 3633098 | 0.296468 |
| Up | DTL | 61 | 0.006526 | 9721978 | 0.297152 |
| Up | CETN2 | 60 | 0.009249 | 16418248 | 0.269586 |
| Up | MCM10 | 59 | 0.007694 | 9107150 | 0.300602 |
| Up | DYNC1I1 | 58 | 0.007347 | 6841812 | 0.299279 |
| Up | PLA2G4A | 57 | 0.006497 | 7102868 | 0.307673 |
| Up | TYMS | 55 | 0.008254 | 2898568 | 0.306378 |
| Up | MDK | 54 | 0.007633 | 2641060 | 0.307322 |
| Up | GOLIM4 | 45 | 0.009056 | 6535396 | 0.299091 |
| Up | TGFB1I1 | 43 | 0.00576 | 2542056 | 0.317363 |
| Up | PKD2 | 42 | 0.006903 | 2134988 | 0.290538 |
| Up | FGA | 41 | 0.004439 | 1801264 | 0.293153 |
| Up | PRKAR1B | 41 | 0.004763 | 4851900 | 0.299424 |
| Up | ACOT7 | 39 | 0.00511 | 6777086 | 0.287968 |
| Up | FGG | 37 | 0.002736 | 1188570 | 0.282125 |
| Up | AP1M2 | 37 | 0.007088 | 9431672 | 0.281894 |
| Up | MITF | 36 | 0.006067 | 5049390 | 0.308118 |
| Up | SMYD3 | 35 | 0.006122 | 1971838 | 0.303776 |
| Up | ACTR3B | 35 | 0.002767 | 5762182 | 0.273907 |
| Up | CTPS2 | 34 | 0.004516 | 1656234 | 0.297352 |
| Up | HP | 33 | 0.004002 | 2476788 | 0.299743 |
| Up | MYL6B | 33 | 0.001721 | 1494552 | 0.303835 |
| Up | LAMB2 | 33 | 0.003266 | 3787290 | 0.283015 |
| Up | ME1 | 32 | 0.00497 | 5458746 | 0.266142 |
| Up | FKBP1B | 31 | 0.002492 | 3149134 | 0.263535 |
| Up | EAF2 | 31 | 0.003666 | 3614748 | 0.273181 |
| Up | CD63 | 30 | 0.002963 | 1542292 | 0.278807 |
| Up | TMEFF1 | 29 | 0.004306 | 1459454 | 0.277382 |
| Up | PHACTR3 | 29 | 0.003235 | 6793662 | 0.273205 |
| Up | MMP1 | 28 | 0.004131 | 1062610 | 0.285972 |
| Up | RAB32 | 28 | 0.004125 | 3089688 | 0.273326 |
| Up | NT5DC2 | 27 | 0.003149 | 4016356 | 0.290497 |
| Up | LAPTM4B | 26 | 0.003861 | 1308630 | 0.280985 |
| Up | LCN2 | 26 | 0.003047 | 3293226 | 0.261121 |
| Up | GRB14 | 26 | 0.001737 | 2102726 | 0.272171 |
| Up | PRTFDC1 | 26 | 0.003559 | 2778424 | 0.248364 |
| Up | SERPINE2 | 25 | 0.003606 | 2767974 | 0.266199 |
| Up | APOA2 | 24 | 0.003639 | 1777538 | 0.282035 |
| Up | CNN1 | 24 | 0.002696 | 1398988 | 0.281984 |
| Up | MAGI2 | 24 | 0.003636 | 4072986 | 0.273277 |
| Up | PADI4 | 24 | 0.001853 | 2786250 | 0.285457 |
| Up | ARNTL2 | 23 | 0.004537 | 2736046 | 0.276156 |
| Up | MAL2 | 23 | 0.004035 | 3057928 | 0.266555 |
| Up | TAX1BP3 | 22 | 0.00296 | 2483404 | 0.27773 |
| Up | APOH | 22 | 0.002261 | 1011952 | 0.294759 |
| Up | GLA | 22 | 0.002185 | 2812520 | 0.25131 |
| Up | KRT7 | 22 | 0.002247 | 3354252 | 0.283132 |
| Up | ROBO1 | 22 | 0.002997 | 2829202 | 0.285181 |
| Up | ADAMTS1 | 21 | 0.003523 | 2991386 | 0.249044 |
| Up | OXTR | 21 | 0.003005 | 1885024 | 0.254274 |
| Up | ADAM32 | 21 | 0.002751 | 2226220 | 0.244362 |
| Up | TSPAN15 | 20 | 0.002903 | 1636154 | 0.248684 |
| Up | DMC1 | 20 | 0.001518 | 1667540 | 0.255218 |
| Up | EPDR1 | 20 | 0.00263 | 955076 | 0.289735 |
| Up | LACTB2 | 20 | 0.003285 | 3861294 | 0.241888 |
| Up | UNC13B | 19 | 0.003409 | 6715552 | 0.228252 |
| Up | ERC2 | 19 | 0.002956 | 2368572 | 0.246592 |
| Up | HRG | 19 | 0.002437 | 1023636 | 0.273991 |
| Up | SMPD1 | 19 | 0.00236 | 2063048 | 0.275125 |
| Up | ZFPM2 | 19 | 0.001198 | 1822000 | 0.266153 |
| Up | METTL22 | 19 | 0.001598 | 2257038 | 0.269023 |
| Up | ORM1 | 18 | 0.001349 | 1072266 | 0.255461 |
| Up | ZC3HAV1L | 18 | 0.001054 | 1624786 | 0.281202 |
| Up | STAC | 18 | 0.001798 | 2388538 | 0.275578 |
| Up | POLR3G | 17 | 0.003237 | 9664174 | 0.229055 |
| Up | DDAH1 | 17 | 0.003704 | 1699018 | 0.290824 |
| Up | TIMP1 | 17 | 0.002047 | 1211820 | 0.264888 |
| Up | DERA | 17 | 0.001512 | 1709944 | 0.240983 |
| Up | TGFB3 | 16 | 0.001732 | 1135010 | 0.26056 |
| Up | ANXA3 | 16 | 0.002306 | 3218636 | 0.263928 |
| Up | ETV4 | 16 | 0.00158 | 1430402 | 0.286767 |
| Up | RND3 | 16 | 0.001618 | 1798784 | 0.267963 |
| Up | FUNDC1 | 16 | 0.001881 | 5031262 | 0.246033 |
| Up | PPP1R14A | 16 | 0.001348 | 1468196 | 0.269398 |
| Up | ADAM22 | 16 | 9.66E-04 | 1467492 | 0.278092 |
| Up | TRHDE | 16 | 0.002417 | 2718244 | 0.265571 |
| Up | AOPEP | 15 | 5.09E-04 | 455628 | 0.281778 |
| Up | LOXL3 | 15 | 0.002245 | 2647672 | 0.272735 |
| Up | TST | 15 | 0.002644 | 2760934 | 0.24058 |
| Up | CENPI | 15 | 6.90E-04 | 213372 | 0.26143 |
| Up | TRPC6 | 15 | 0.002982 | 3195972 | 0.236597 |
| Up | MSANTD3 | 14 | 0.001423 | 599870 | 0.282602 |
| Up | DCBLD2 | 14 | 0.001863 | 1414006 | 0.280806 |
| Up | ICA1 | 14 | 0.001837 | 1818980 | 0.253566 |
| Up | CABLES1 | 14 | 4.42E-04 | 448806 | 0.275222 |
| Up | GTF3C6 | 13 | 0.001446 | 393772 | 0.300413 |
| Up | AVEN | 13 | 7.46E-04 | 364352 | 0.294788 |
| Up | PBLD | 13 | 0.001491 | 1062854 | 0.235839 |
| Up | CD151 | 13 | 0.001883 | 1879220 | 0.264662 |
| Up | YIF1B | 13 | 0.001267 | 1419634 | 0.263917 |
| Up | C5orf30 | 13 | 0.001783 | 1452154 | 0.265582 |
| Up | FRMD3 | 11 | 8.55E-04 | 686546 | 0.235579 |
| Up | FAH | 11 | 0.001509 | 1294972 | 0.259587 |
| Up | HTATIP2 | 11 | 0.00148 | 1272602 | 0.261231 |
| Up | CYSTM1 | 11 | 0.002293 | 1438306 | 0.2152 |
| Up | MAP1LC3B2 | 10 | 0.001139 | 1052378 | 0.240608 |
| Up | MYEOV | 2 | 5.45E-05 | 24642 | 0.227079 |
| Up | DNAH14 | 1 | 0 | 0 | 0.212349 |
| Up | MYOM1 | 1 | 0 | 0 | 0.247539 |
| Down | HEPACAM2 | 496 | 0.004088 | 4614336 | 0.259565 |
| Down | HLA-DPA1 | 443 | 0.021345 | 6656236 | 0.294395 |
| Down | DCAF12 | 359 | 0.002195 | 1838728 | 0.28738 |
| Down | POLM | 259 | 6.45E-05 | 60748 | 0.246809 |
| Down | TUBB1 | 231 | 0.017014 | 13445244 | 0.31686 |
| Down | KMT2D | 211 | 0.004121 | 6143304 | 0.276119 |
| Down | TNFRSF13B | 208 | 0.004558 | 7473486 | 0.25441 |
| Down | SEC16A | 199 | 0.024345 | 23456762 | 0.332778 |
| Down | AHNAK | 158 | 0.012868 | 12992410 | 0.339572 |
| Down | OGT | 151 | 0.02538 | 23785672 | 0.321486 |
| Down | TRAK2 | 148 | 0.005447 | 1889278 | 0.291359 |
| Down | PER1 | 143 | 0.007889 | 3343992 | 0.297667 |
| Down | TXNIP | 142 | 0.005831 | 3723272 | 0.3011 |
| Down | GPRASP1 | 140 | 0.011046 | 5269570 | 0.284316 |
| Down | CTSB | 139 | 0.011516 | 5746824 | 0.316973 |
| Down | ITGA4 | 132 | 0.110358 | 99169576 | 0.373747 |
| Down | CBLB | 131 | 0.011859 | 9651222 | 0.31117 |
| Down | LEF1 | 111 | 0.008924 | 3863000 | 0.305275 |
| Down | PLEC | 110 | 0.025923 | 21209448 | 0.346999 |
| Down | BNIP3L | 110 | 0.005843 | 2394548 | 0.292184 |
| Down | CAMK1D | 109 | 0.007993 | 5854240 | 0.294255 |
| Down | NR4A1 | 99 | 0.024611 | 10885038 | 0.333639 |
| Down | OPA1 | 94 | 0.007387 | 5152054 | 0.317804 |
| Down | EEF2 | 83 | 0.037887 | 47075150 | 0.354698 |
| Down | SEMA7A | 82 | 0.00308 | 4153990 | 0.271146 |
| Down | RPS6KA5 | 78 | 0.010004 | 5767566 | 0.321653 |
| Down | STK17B | 78 | 0.00446 | 1356036 | 0.284918 |
| Down | P2RX5 | 75 | 0.003754 | 3579124 | 0.255008 |
| Down | SLC2A1 | 68 | 0.003791 | 2359116 | 0.318622 |
| Down | GYPB | 66 | 0.007619 | 6671358 | 0.262863 |
| Down | HBB | 65 | 0.01032 | 10858208 | 0.304059 |
| Down | CD22 | 64 | 0.001809 | 1124092 | 0.279752 |
| Down | CD4 | 64 | 0.018753 | 14195846 | 0.322845 |
| Down | CD44 | 63 | 0.031474 | 16859342 | 0.344469 |
| Down | TFRC | 63 | 0.019104 | 17774714 | 0.337221 |
| Down | PTPRC | 62 | 0.012352 | 6796924 | 0.316681 |
| Down | CD5 | 62 | 0.00189 | 1129318 | 0.283456 |
| Down | HLA-DRA | 61 | 0.009348 | 4428892 | 0.287995 |
| Down | HLA-DMB | 60 | 0.002503 | 1238774 | 0.277344 |
| Down | CELSR1 | 60 | 0.003079 | 1745004 | 0.272928 |
| Down | MS4A1 | 58 | 0.0017 | 1236694 | 0.276378 |
| Down | SPTA1 | 55 | 0.007158 | 4511084 | 0.30275 |
| Down | IL7R | 54 | 0.027811 | 45141856 | 0.327043 |
| Down | GYPA | 53 | 8.14E-04 | 390278 | 0.260045 |
| Down | SLC4A1 | 53 | 0.002142 | 1341058 | 0.273169 |
| Down | EPB42 | 52 | 0.002323 | 1787000 | 0.260879 |
| Down | RHAG | 49 | 0 | 0 | 0.214566 |
| Down | CD79A | 49 | 0.008789 | 4491848 | 0.293056 |
| Down | CD6 | 48 | 0 | 0 | 0.220862 |
| Down | CSF1R | 48 | 0.002723 | 5503564 | 0.281868 |
| Down | IKZF3 | 48 | 0.021447 | 15624140 | 0.316487 |
| Down | POU2F2 | 46 | 0.002814 | 4242740 | 0.262773 |
| Down | IGF2R | 45 | 0.005336 | 8354606 | 0.300049 |
| Down | NSUN3 | 45 | 0 | 0 | 0.226647 |
| Down | BLK | 44 | 0.009001 | 4827382 | 0.309644 |
| Down | VCAN | 44 | 0.00605 | 2854094 | 0.288546 |
| Down | ADAM28 | 41 | 0 | 0 | 0.272076 |
| Down | RPL27A | 41 | 0.009338 | 13186584 | 0.337921 |
| Down | RPL23A | 39 | 0.022449 | 25286012 | 0.35326 |
| Down | EEF1A1 | 37 | 0.098882 | 1.23E+08 | 0.370457 |
| Down | RPL13A | 37 | 0.009659 | 13645424 | 0.339386 |
| Down | RPL10 | 35 | 0.066934 | 69760500 | 0.352275 |
| Down | NOTCH2 | 35 | 0.009449 | 4095454 | 0.307291 |
| Down | ATP2B1 | 35 | 0.00508 | 5650696 | 0.303835 |
| Down | SLC38A1 | 35 | 0.008346 | 5880382 | 0.292709 |
| Down | QSOX2 | 35 | 0.00527 | 3561792 | 0.291167 |
| Down | SORL1 | 34 | 0.01017 | 6644994 | 0.299569 |
| Down | LRP1 | 34 | 0.030447 | 29146796 | 0.321269 |
| Down | SRRM2 | 34 | 0.044661 | 52105132 | 0.347662 |
| Down | ABCC13 | 34 | 0 | 0 | 0.250182 |
| Down | ATM | 33 | 0.040568 | 31808618 | 0.330997 |
| Down | FECH | 33 | 0.007383 | 3340210 | 0.299511 |
| Down | CD27 | 32 | 0.004882 | 2960412 | 0.278142 |
| Down | VIPR1 | 32 | 0.004948 | 5472644 | 0.278694 |
| Down | AGPAT4 | 32 | 1.70E-05 | 28760 | 0.230987 |
| Down | CIITA | 32 | 0.003427 | 3117716 | 0.289328 |
| Down | EP400 | 31 | 0.007237 | 9601258 | 0.288196 |
| Down | RORA | 31 | 0.004548 | 1396286 | 0.302558 |
| Down | SOX6 | 31 | 0.001788 | 1768940 | 0.255661 |
| Down | CENPF | 30 | 0.008068 | 6644598 | 0.297681 |
| Down | FAM167A | 29 | 0.008587 | 3540932 | 0.276168 |
| Down | MXI1 | 28 | 0.001985 | 3293278 | 0.269504 |
| Down | ALDH5A1 | 27 | 0.003494 | 4468018 | 0.270813 |
| Down | RPL37 | 27 | 0.001913 | 3460916 | 0.29617 |
| Down | TSPAN5 | 26 | 0.01221 | 9998796 | 0.2873 |
| Down | PIEZO1 | 26 | 0 | 0 | 0.223188 |
| Down | BTG1 | 25 | 0.002334 | 3106952 | 0.271777 |
| Down | SPIB | 25 | 0.001073 | 679476 | 0.280679 |
| Down | ALDH6A1 | 25 | 0.00345 | 3839804 | 0.246053 |
| Down | COL7A1 | 25 | 0.003621 | 3830672 | 0.277705 |
| Down | PAX5 | 25 | 0.008408 | 15230588 | 0.288317 |
| Down | DUSP2 | 24 | 0.003694 | 4774362 | 0.262573 |
| Down | IFFO1 | 24 | 0.004108 | 2648480 | 0.262006 |
| Down | COBLL1 | 24 | 0.00318 | 3800612 | 0.267373 |
| Down | DBP | 24 | 0.00207 | 2593884 | 0.258708 |
| Down | SELENBP1 | 24 | 0.011965 | 15173332 | 0.275259 |
| Down | DYRK2 | 23 | 0.00992 | 5736550 | 0.313918 |
| Down | PDE3B | 23 | 0.005858 | 5188848 | 0.268242 |
| Down | ABCA2 | 22 | 0.002246 | 1887488 | 0.270647 |
| Down | TTN | 22 | 0.022803 | 22179002 | 0.328956 |
| Down | AAK1 | 22 | 0.001768 | 1776702 | 0.306636 |
| Down | CAMK4 | 21 | 0.001804 | 1122842 | 0.282382 |
| Down | ANKRD52 | 21 | 0.004706 | 3610412 | 0.290251 |
| Down | OBSCN | 21 | 0.002307 | 1424536 | 0.295125 |
| Down | YOD1 | 21 | 0.003727 | 4452540 | 0.278581 |
| Down | RNF213 | 21 | 0.003281 | 4225796 | 0.28073 |
| Down | HBM | 21 | 0.003971 | 2893622 | 0.211891 |
| Down | FAM117B | 20 | 0.002939 | 3060242 | 0.274465 |
| Down | RANBP10 | 20 | 0.010162 | 16110914 | 0.263344 |
| Down | RALGPS2 | 20 | 0.008034 | 10752578 | 0.280386 |
| Down | CNKSR2 | 19 | 0.003716 | 4283878 | 0.253753 |
| Down | MYO15B | 19 | 0.001548 | 1850770 | 0.253763 |
| Down | BMF | 18 | 0.002348 | 3575424 | 0.266842 |
| Down | WDFY2 | 18 | 0.004928 | 3808614 | 0.241237 |
| Down | LENG8 | 18 | 0.006353 | 9944066 | 0.270896 |
| Down | TUBGCP6 | 18 | 0.00405 | 3821222 | 0.278155 |
| Down | NELL2 | 17 | 0.005907 | 5985122 | 0.250435 |
| Down | YIPF4 | 17 | 0.002754 | 3249620 | 0.263928 |
| Down | OSBPL10 | 17 | 0.002471 | 663904 | 0.287727 |
| Down | BACH2 | 16 | 0.002632 | 2910574 | 0.257534 |
| Down | NLRP1 | 16 | 0.003929 | 3755972 | 0.280691 |
| Down | BCL11B | 2 | 0.001184 | 2467486 | 0.258362 |
| Down | STRADB | 2 | 0.001267 | 1783952 | 0.276898 |
| Down | BCL11A | 1 | 0.004017 | 6158386 | 0.273664 |
| Down | RCAN3 | 1 | 0 | 0 | 0.233006 |
| Down | KLHL3 | 1 | 0.003155 | 2811806 | 0.280551 |
| Down | CLEC2D | 1 | 0.015351 | 13153684 | 0.270766 |
| Down | TNRC6B | 1 | 0.010638 | 19725384 | 0.302365 |
| Down | FBXO7 | 1 | 0.011435 | 14645282 | 0.298528 |
| Down | MOB3B | 1 | 0 | 0 | 0.202401 |
| Down | ZNF549 | 1 | 0 | 0 | 0.230523 |
| Down | VSTM2A | 1 | 0 | 0 | 0.220343 |
| Down | CTC1 | 1 | 0 | 0 | 0.200285 |
| Down | MRC2 | 1 | 0.002911 | 3908206 | 0.26787 |
| Down | TTC14 | 1 | 0 | 0 | 0.257985 |
| Down | ADA2 | 1 | 0 | 0 | 0.257985 |

**Fig. 3.**
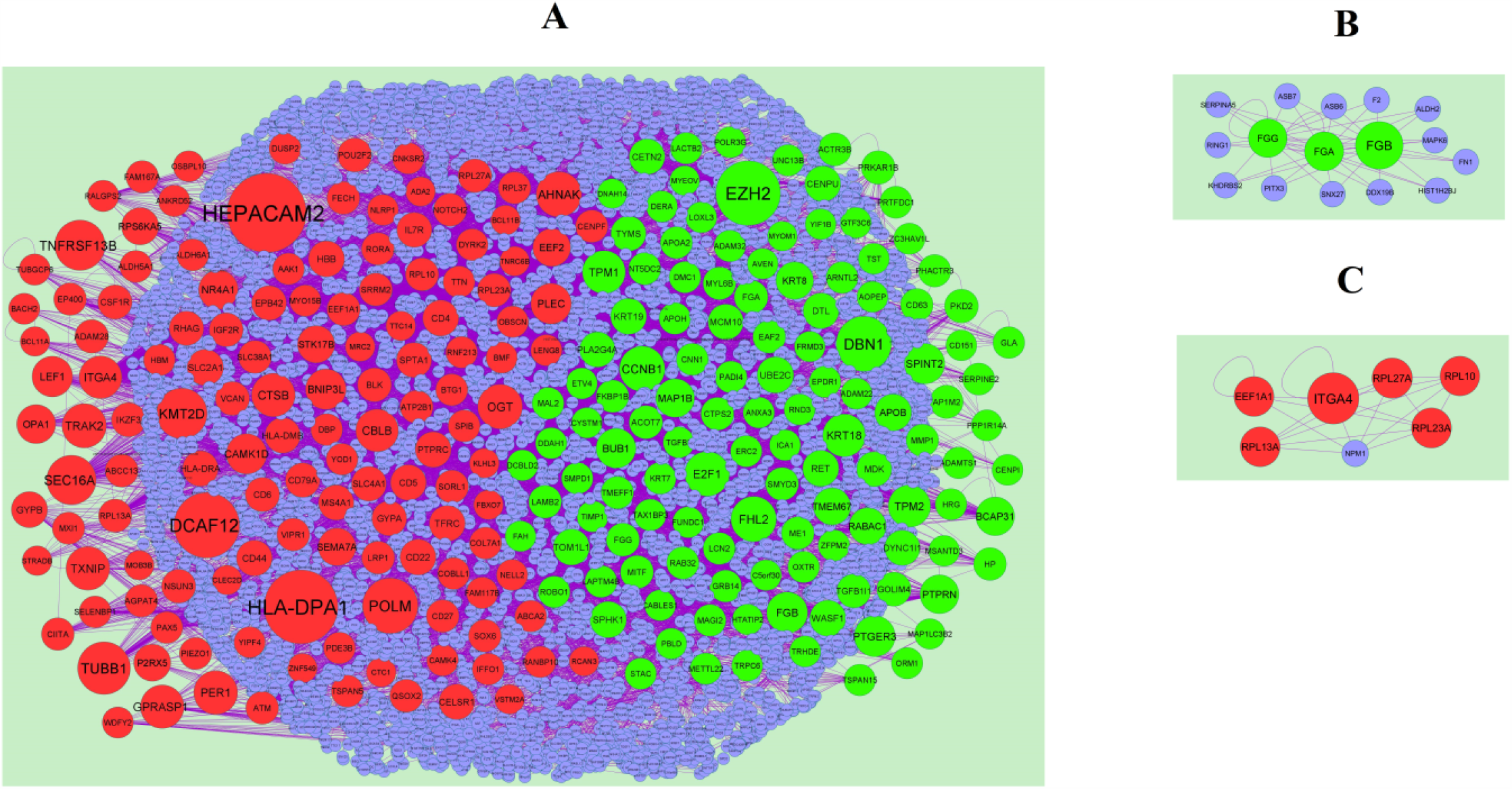
PPI network and the most significant modules of DEGs. (A) The PPI network of DEGs was constructed using Cytoscape (B) The most significant module was obtained from PPI network with 16 nodes and 44 edges for up regulated genes (C) The most significant module was obtained from PPI network with 6 nodes and 20 edges for up regulated genes. Up regulated genes are marked in green; down regulated genes are marked in red

### Construction of miRNA-DEG regulatory network

The regulatory network of miRNA-DEG and predicted targets is presented in Fig. 4A. Notably, MAP1B targeted 202 miRNAs, including hsa-mir-4461; CCNB1 targeted 94 miRNAs, including hsa-mir-3928-3p; AHNAK targeted 256 miRNAs, including hsa-mir-2682-5p; KMT2D targeted 209 miRNAs, including hsa-mir-1202 and top 20 are listed in Table 6. As a group, a total of 257 of the 463 DEGs were contained in the miRNA-DEG regulatory network.

**Table 6.** miRNA - target gene and TF - target gene interaction

| Regulation | Target Genes | Degree | MicroRNA | Regulation | Target Genes | Degree | TF |
| --- | --- | --- | --- | --- | --- | --- | --- |
| Up | COL1A1 | 178 | hsa-mir-4492 | Up | IRF4 | 10 | NFATC2 |
| Up | IRF4 | 140 | hsa-mir-4319 | Up | LCK | 10 | YY1 |
| Up | MYBL2 | 83 | hsa-mir-637 | Up | RET | 10 | NR2C2 |
| Up | PRKCB | 81 | hsa-mir-1261 | Up | MAP1LC3C | 10 | MAX |
| Up | IL2RB | 54 | hsa-mir-4300 | Up | IL2RB | 8 | PDX1 |
| Up | CCR5 | 50 | hsa-mir-5193 | Up | GRAP2 | 8 | ELK1 |
| Up | GRAP2 | 41 | hsa-mir-3681-5p | Up | MYBPC2 | 7 | RELA |
| Up | MDFI | 41 | hsa-mir-4441 | Up | MDFI | 6 | TFAP2A |
| Up | RET | 17 | hsa-mir-129-2-3p | Up | MYBL2 | 6 | GATA2 |
| Up | IKZF1 | 16 | hsa-mir-3607-3p | Up | CD247 | 5 | SREBF1 |
| Up | BTK | 13 | hsa-mir-4667-3p | Up | COL1A1 | 5 | NFYA |
| Up | LCK | 6 | hsa-mir-210-3p | Up | PRKCB | 4 | IRF2 |
| Up | MYBPC2 | 6 | hsa-mir-214-3p | Up | IKZF1 | 4 | E2F6 |
| Up | CD247 | 4 | hsa-mir-346 | Up | BTK | 2 | SOX5 |
| Up | MAP1LC3C | 2 | hsa-mir-27a-3p | Up | CCR5 | 1 | EGR1 |
| Down | JUN | 144 | hsa-mir-3943 | Down | ATF3 | 19 | TP53 |
| Down | EGR1 | 132 | hsa-mir-548e-3p | Down | EGR1 | 16 | ARID3A |
| Down | ZFP36 | 130 | hsa-mir-6077 | Down | JUNB | 15 | SRF |
| Down | FOS | 105 | hsa-mir-5586-5p | Down | FOS | 13 | CREB1 |
| Down | DUSP1 | 97 | hsa-mir-4458 | Down | PTPRO | 12 | NR3C1 |
| Down | JUNB | 85 | hsa-mir-3065-5p | Down | NR0B2 | 11 | USF1 |
| Down | MME | 54 | hsa-mir-922 | Down | MME | 9 | BRCA1 |
| Down | NR4A2 | 50 | hsa-mir-29b-2-5p | Down | JUN | 9 | SP1 |
| Down | ATF3 | 48 | hsa-mir-5000-5p | Down | DUSP1 | 9 | STAT3 |
| Down | NR4A1 | 43 | hsa-mir-107 | Down | NR4A1 | 9 | HINFP |
| Down | PCK1 | 38 | hsa-mir-1185-1-3p | Down | NR4A2 | 7 | NR2E3 |
| Down | PTPRO | 18 | hsa-mir-203a-3p | Down | PCK1 | 6 | NR2F1 |
| Down | APOB | 17 | hsa-mir-548p | Down | ZFP36 | 5 | TFAP2C |
| Down | ALB | 10 | hsa-mir-492 | Down | APOB | 4 | FOXA1 |
| Down | NR0B2 | 5 | hsa-mir-141-3p | Down | ALB | 4 | STAT1 |

**Fig. 4.**
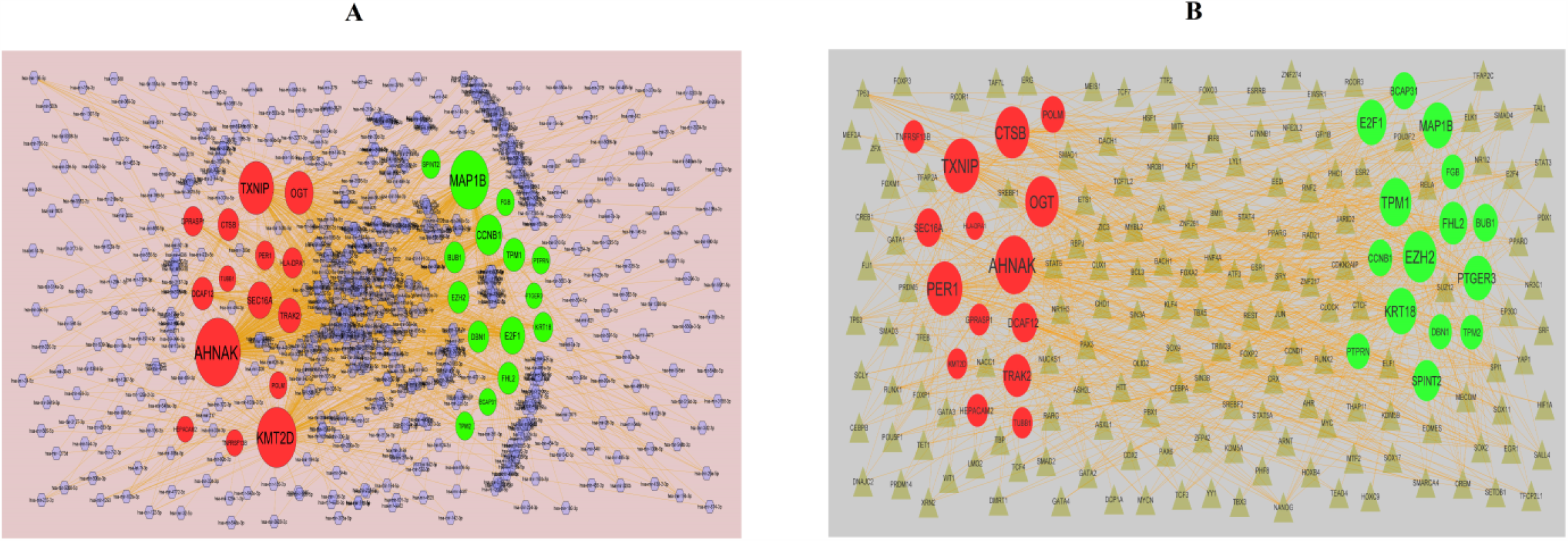
(A) Target gene - miRNA regulatory network between target genes and miRNAs (B) Target gene - TF regulatory network between target genes and TFs. Up regulated genes are marked in green; down regulated genes are marked in red; The blue color diamond nodes represent the key miRNAs; the gray color triangle nodes represent the key TFs

### Construction of TF-DEG regulatory network

The regulatory network of TF-DEG and predicted targets is presented in Fig. 4B. Notably, EZH2 targeted 45 TFs, including SOX2; TPM1 targeted 40 TFs, including MYC; AHNAK targeted 58 TFs, including KLF4, TXNIP targeted 51 TFs, including TP63 and top 20 are listed in Table 6. As a group, a total of 259 of the 463 DEGs were contained in the TF-DEG regulatory network.

### Hub genes validation

All of the hub genes were validated in TCGA data. Hub genes contributed to the survival period in patients with PDAC, we analyzed the overall survival (OS) for each hub gene by UALCAN (Fig. 5). The results showed that the high expression of CCNB1 and FHL2 mRNA level were associated with the worse OS in patients with PDAC, while low expression of HLA-DPA1 and TUBB1 mRNA level were associated with the worse OS in patients with PDAC. As shown in Fig. 6, the expression of the up regulated hub genes CCNB1 and FHL2 in PDAC were significantly elevated compared with normal, while expression of the down regulated hub genes HLA-DPA1 and TUBB1 in PDAC were significantly decreased compared with normal. The expression of each hub gene in PDAC patients was analyzed according to the individual cancer stage. As shown in Fig. 7, the expression of CCNB1 and FHL2 were higher in patients with all individual cancer stages than that in normal, which revealed that these up regulated hub genes might be associated with tumor progression positively, where as the expression of HLA-DPA1 and TUBB1 were lower in patients with all individual cancer stages than that in normal, which revealed that these down regulated hub genes might be associated with tumor progression positively. We used cBioportal tool to explore the specific mutation of hub genes in PDAC dataset with 184 samples. From the OncoPrint, percentages of alterations in CCNB1, FHL2, HLA-DPA1 and TUBB1 genes among lung cancer ranged from 0% to 2.3% in individual genes (CCNB1, 0%; FHL2, 0.6%; HLA-DPA1, 2.3%; TUBB1, 2.3%) and is shown in Fig. 8. In addition, we used the ‘HPA’ to examine the protein expression levels of CCNB1 and FHL2, and observed that the protein expression levels of the these hub genes were noticeably up regulated in PDAC compared with normal tissues, whereas protein expression levels of HLA-DPA1 and TUBB1, and observed that the protein expression levels of the these hub genes were noticeably down regulated in PDAC compared with normal tissues (Fig. 9). The association of CCNB1, FHL2, HLA-DPA1 and TUBB1 expression level with immune infiltration abundance in PDAC was evaluated using TIMER database. CCNB1 and FHL2 expression were negatively correlated with infiltration degree of B cells, CD8+ T cells, macrophage, neutrophil, and dendritic cells, where as HLA-DPA1 and TUBB1 were positively correlated with infiltration degree of B cells, CD8+ T cells, macrophage, neutrophil, and dendritic cells and is shown in Fig. 10. As these 4 genes are prominently expressed in PDAC, we performed a ROC curve analysis to evaluate their sensitivity and specificity for the diagnosis of PDAC. As shown in Fig. 11, CCNB1, FHL2, HLA-DPA1 and TUBB1 achieved an AUC value of >0.70, demonstrating that these genes have high sensitivity and specificity for PDAC diagnosis. The results suggested that CCNB1, FHL2, HLA-DPA1 and TUBB1 can be used as biomarkers for the diagnosis of PDAC.

**Fig. 5.**
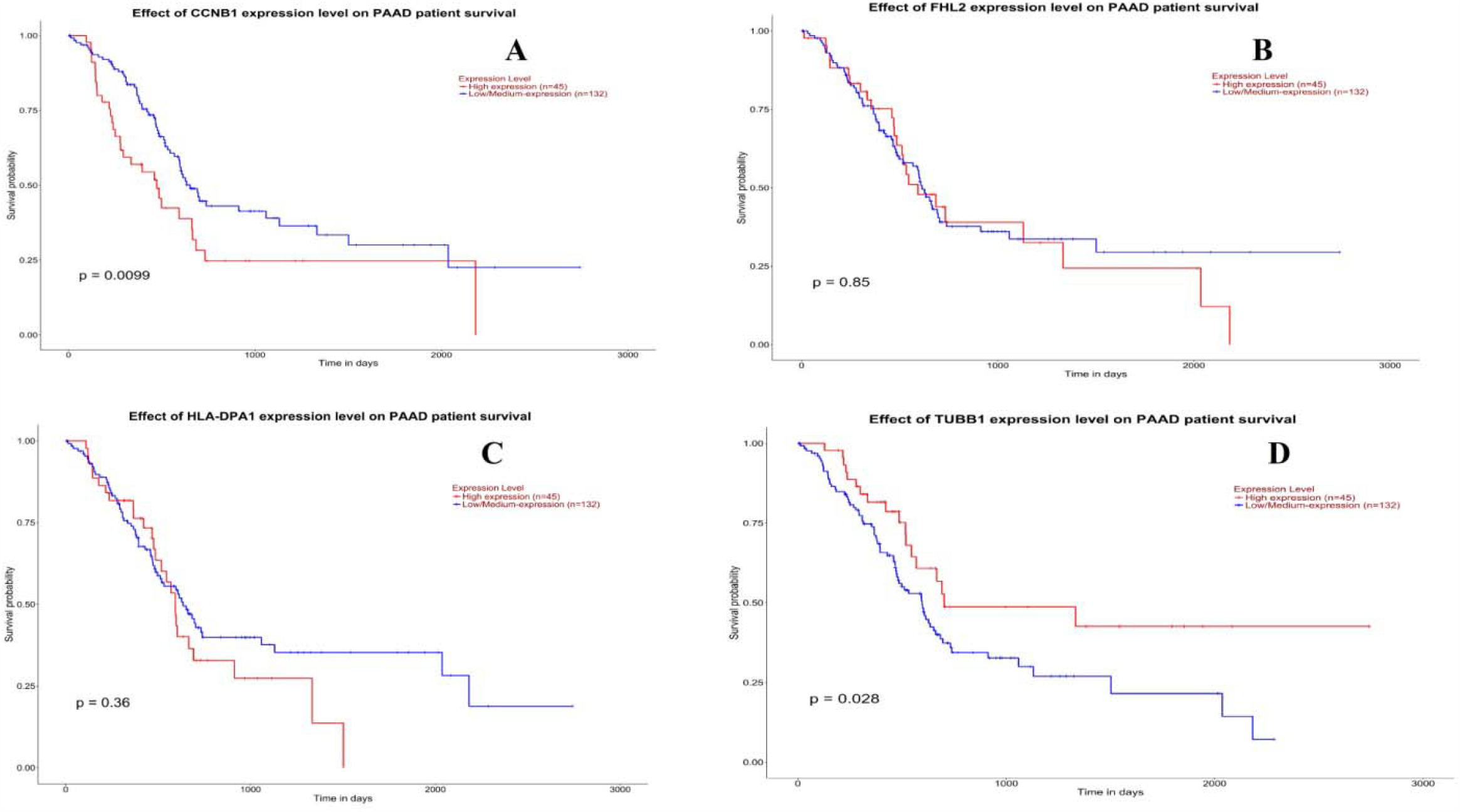
Overall survival analysis of hub genes. Overall survival analyses were performed using the UALCAN online platform. Red line denotes - high expression; Blur line denotes – low expression. A) CCNB1 B) FHL2 C) HLA-DPA1 D) TUBB1

**Fig. 6.**
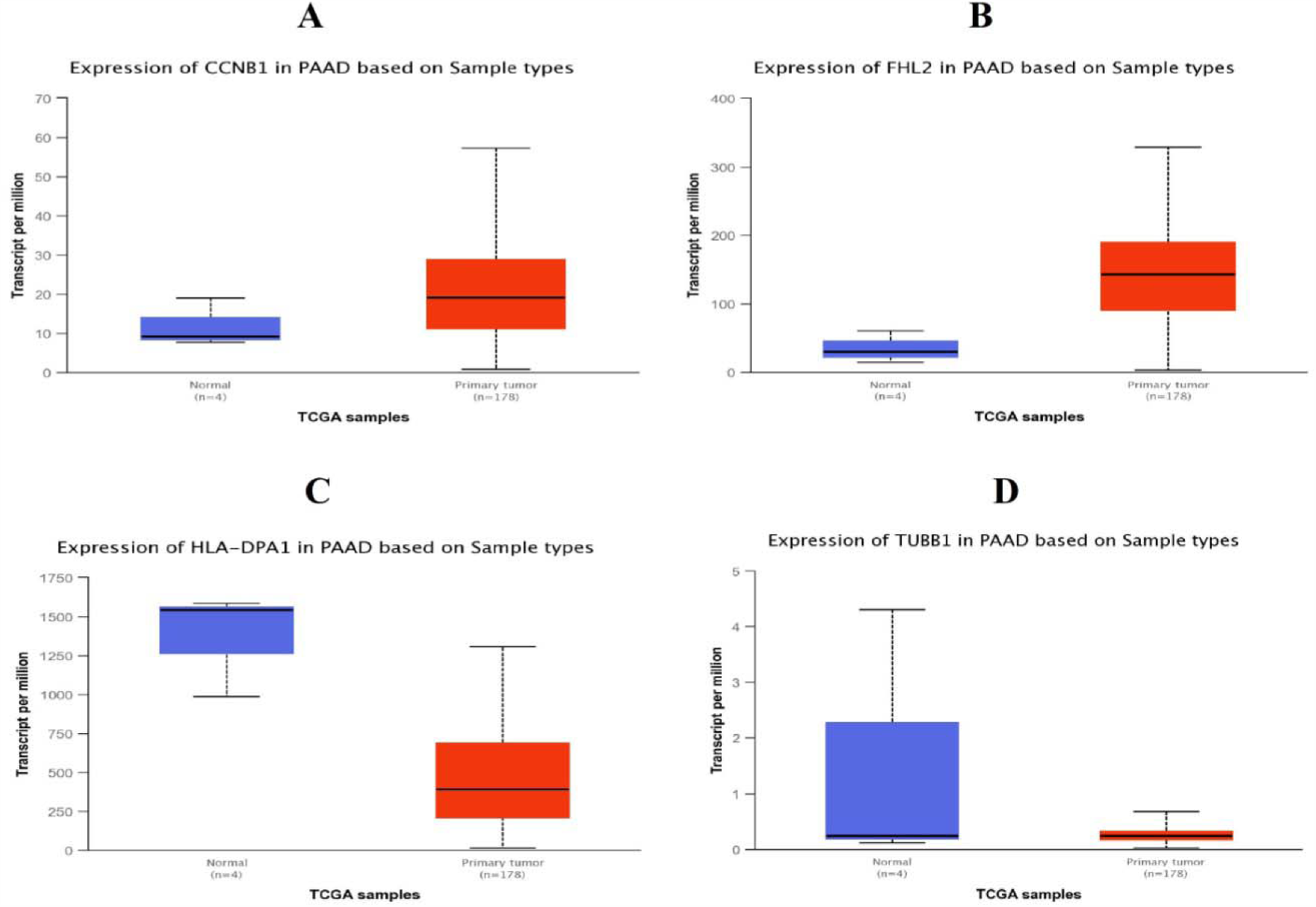
Box plots (expression analysis) hub genes were produced using the UALCAN platform. A) CCNB1 B) FHL2 C) HLA-DPA1 D) TUBB1

**Fig. 7.**
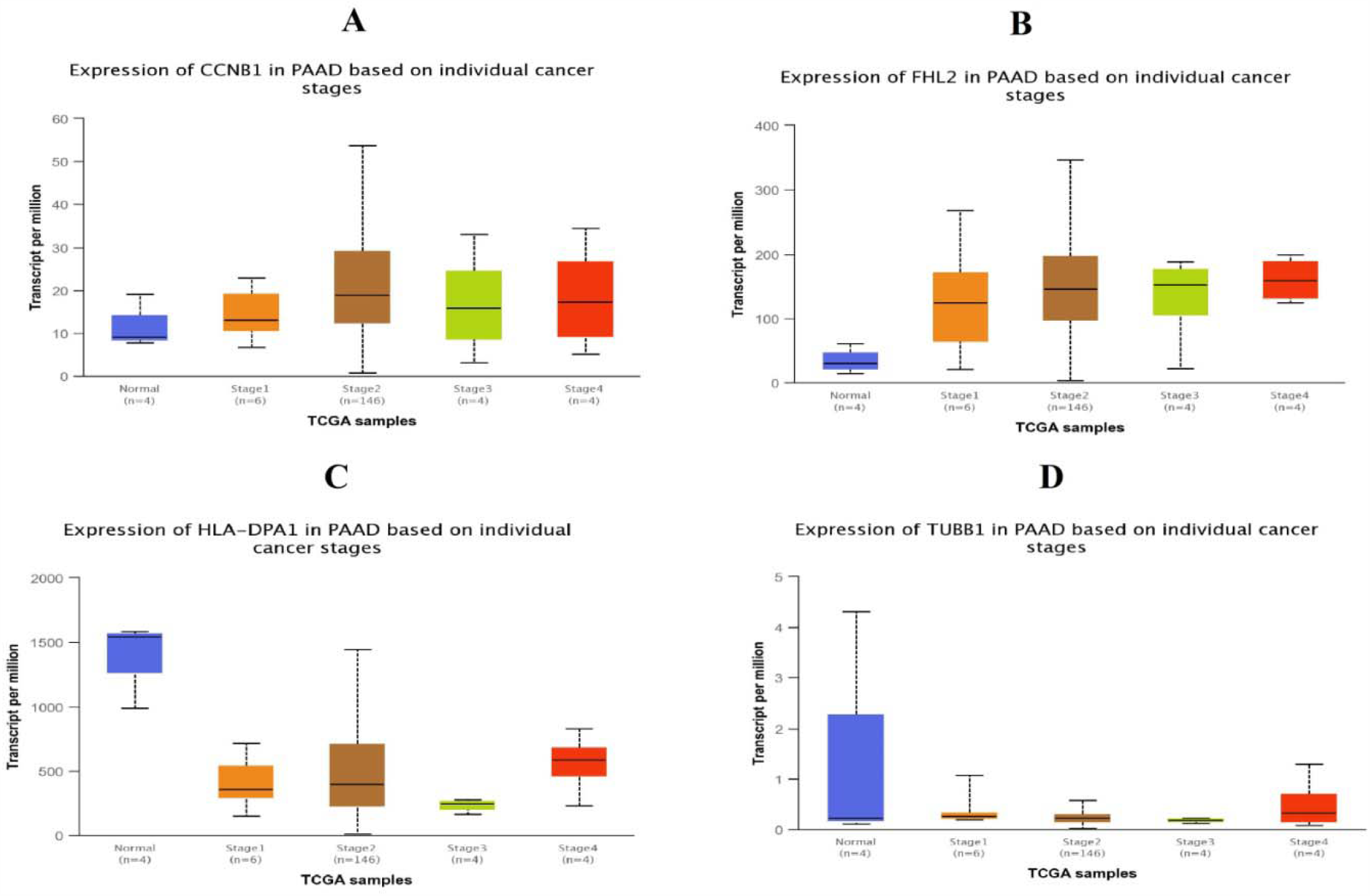
Box plots (clinical stage analysis) hub genes were produced using the UALCAN platform. A) CCNB1 B) FHL2 C) HLA-DPA1 D) TUBB1

**Fig. 8.**
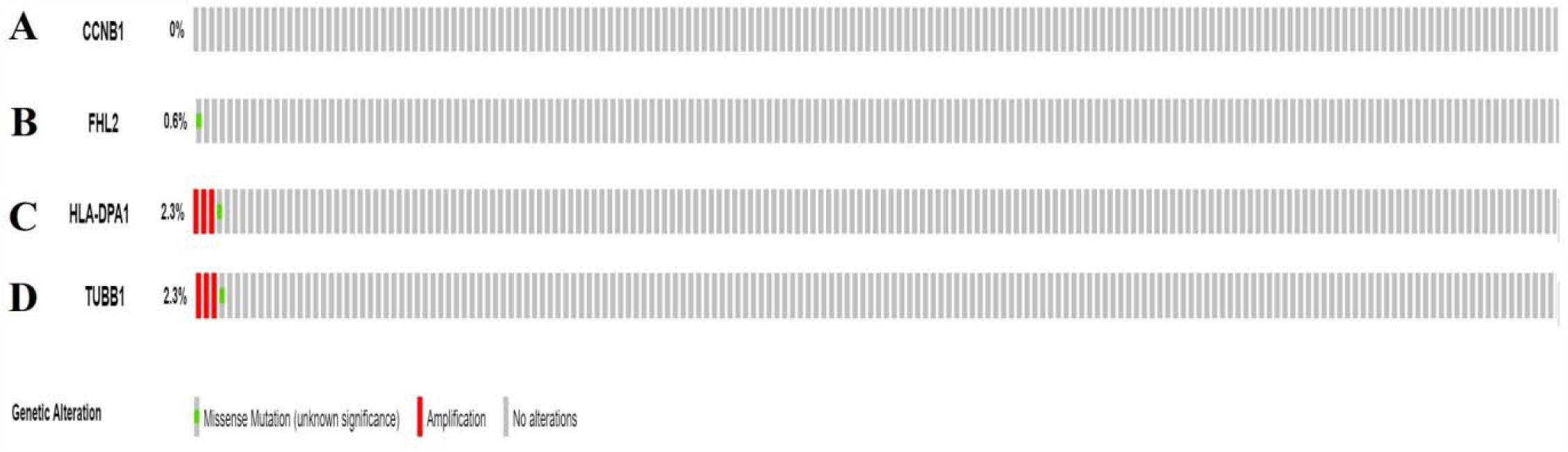
Mutation analyses of hub genes were produced using the CbioPortal online platform. A) CCNB1 B) FHL2 C) HLA-DPA1 D) TUBB1

**Fig. 9.**
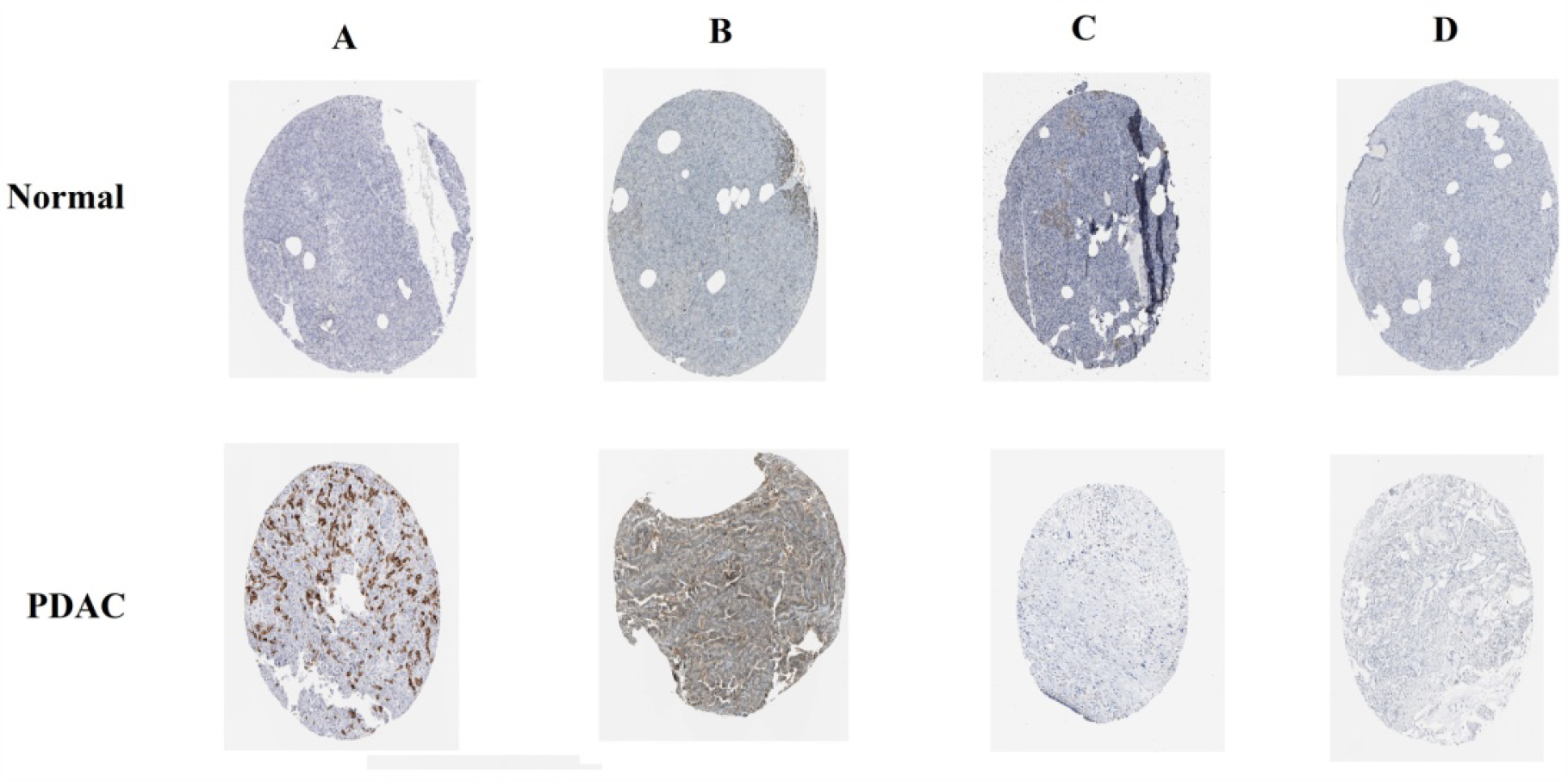
Immunohisto chemical (IHC) analyses of hub genes were produced using the human protein atlas (HPA) online platform. A) CCNB1 B) FHL2 C) HLA-DPA1 D) TUBB1

**Fig. 10.**
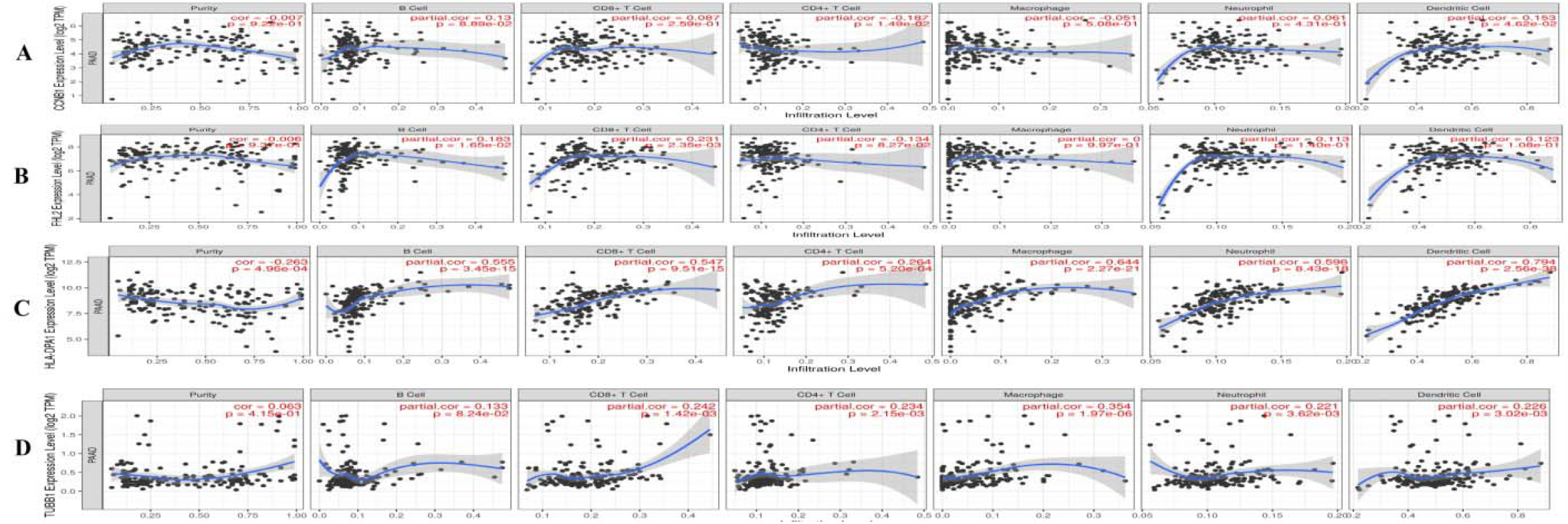
Scatter plot for immune infiltration for hub genes. A) CCNB1 B) FHL2 C) HLA-DPA1 D) TUBB1

**Fig. 11.**
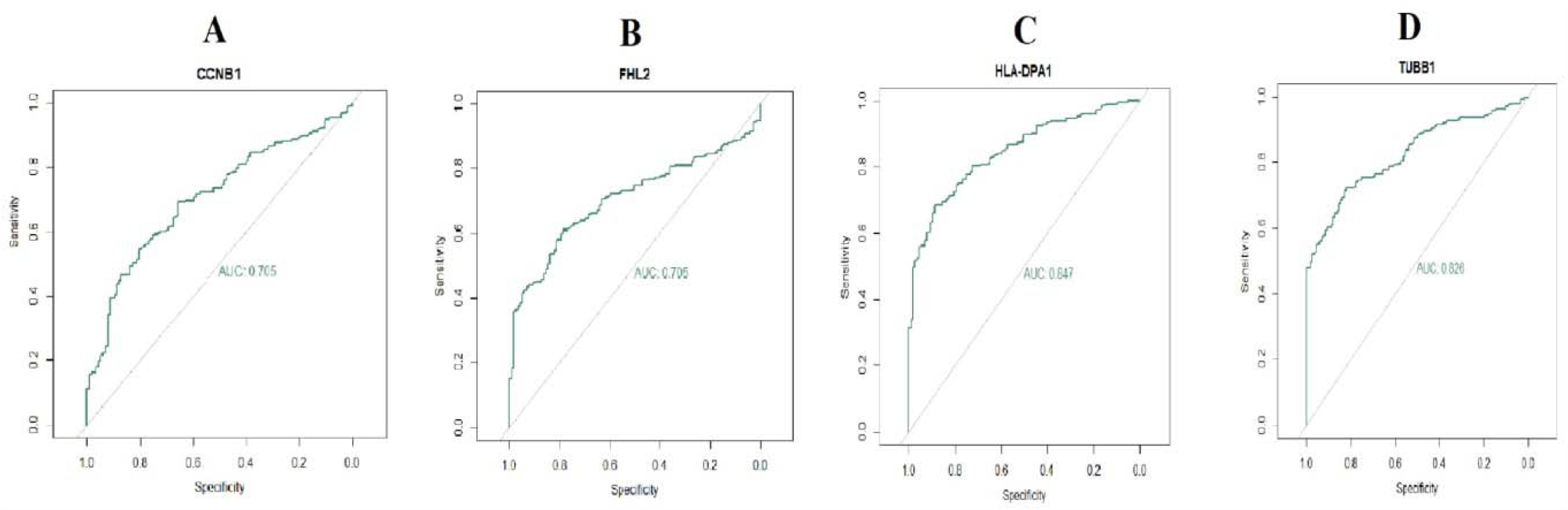
ROC curve validated the sensitivity, specificity of hub genes as a predictive biomarker for PDAC prognosis. A) CCNB1 B) FHL2 C) HLA-DPA1 D) TUBB1

### RT-PCR analysis

Next, in order to verify the results of previous bioinformatics analysis, the gene expression levels of CCNB1, FHL2, HLA-DPA1 and TUBB1 were detected by RT-PCR. As shown in Fig 12, CCNB1 and FHL2 mRNA expression levels were significantly up regulated in the PDAC compared to normal, and HLA-DPA1 and TUBB1 mRNA level were down regulated compared to normal, which was consistent with the results of bioinformatics analysis.

**Fig. 12.**
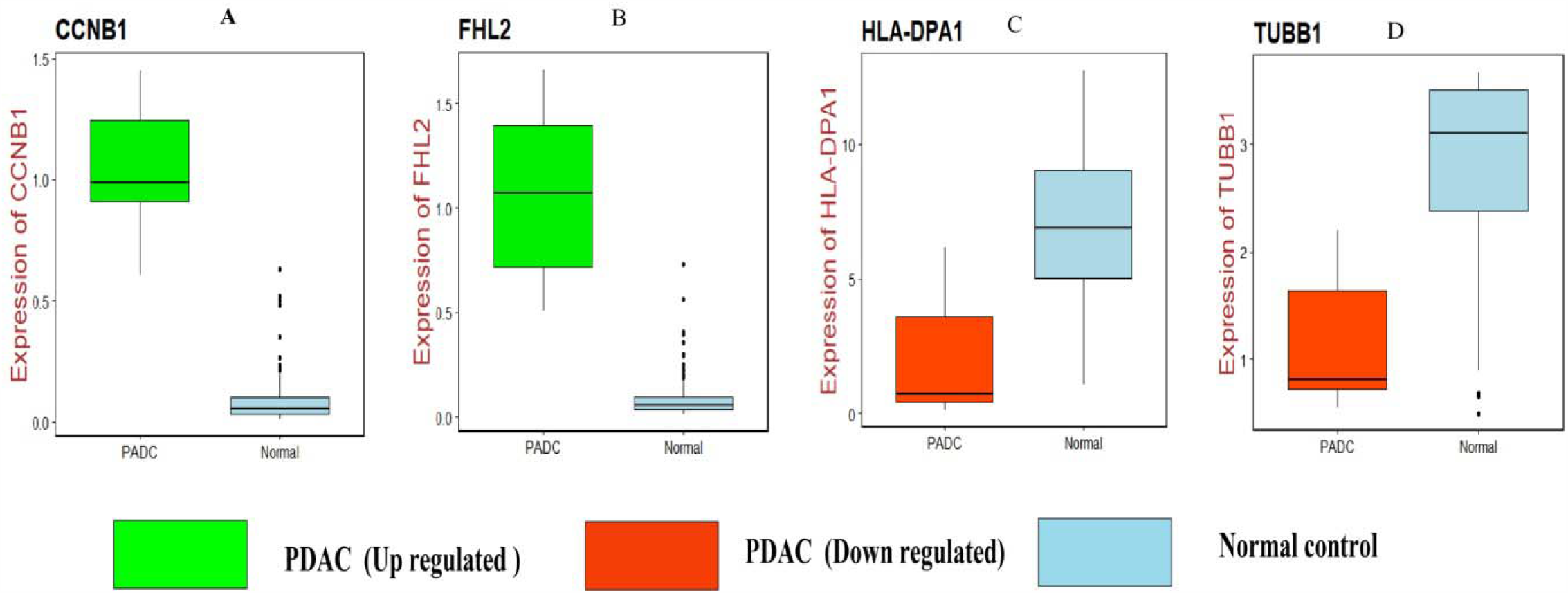
Validation of hub genes by RT-PCR. A) CCNB1 B) FHL2 C) HLA-DPA1 D) TUBB1

### Molecular docking experiment

The docking simulation was performed in the present study to recognize the active site conformation andsignificant interactions, which are responsible for complex stability with ligand receptor. Novel molecules containing alkylating group and purine heterocyclic ring were designed and performed docking studies using Sybyl X 2.1 drug design software. Molecules containing alkylating group is designed due to non-specific alkylation of physiologically important groupings and purine heterocyclic ring is incorporated due to structural similarity of purine derivatives and to compete for the synthesis of proteins. The proteins which are over expressed in pancreatic duct adenocarcinoma are selected for docking studies. The two proteins of each over expressedcyclin B1 (CCNB1)its co-crystallised protein of PDB code 4Y72, 5H0V and Four and half LIM domains 2 (FHL2) of NMR structure of proteins 2D8Z and 2EHE were selected for docking. The investigation of designed molecules was performed to identify the potential molecule. The most of the designed molecules obtained C-score greater than 5 and are active having the c-score greater than 5 are said to be an active, among total of 48 designed molecules few molecules have excellent good binding energy (C-score) greater than 8 respectively. Few of the designed molecules IM 11 & PU 42, shown good binding score of 7.83& 8.57 and the molecules IM 13, TZ 23, TZ 27, TZ 37, PU 41, PU 43 & PU 49 have good binding score 8.013, 8.523, 8.235, 8.800, 10.338, 10.891& 9.411 with CCNB1 PDB code 1H0V and 4Y72 respectively and are shown in Fig. 13. Molecules of IM 09, IM 10, IM & 18 shown good binding score of 7.14, 7.75 & 7.80 and the molecules IM 8 and TZ 24 with binding score 6.24 and 6.32 with FHL2 of PDB code 2D8Z and 2EQQ respectively, the values are depicted in Table 7. The molecule IM 8has highest binding score its interaction with protein 1H0V and hydrogen bonding and other bonding interactions with amino acids are depicted by 3D and 2D shown in Fig. 14 and Fig. 15.

**Table 7.**
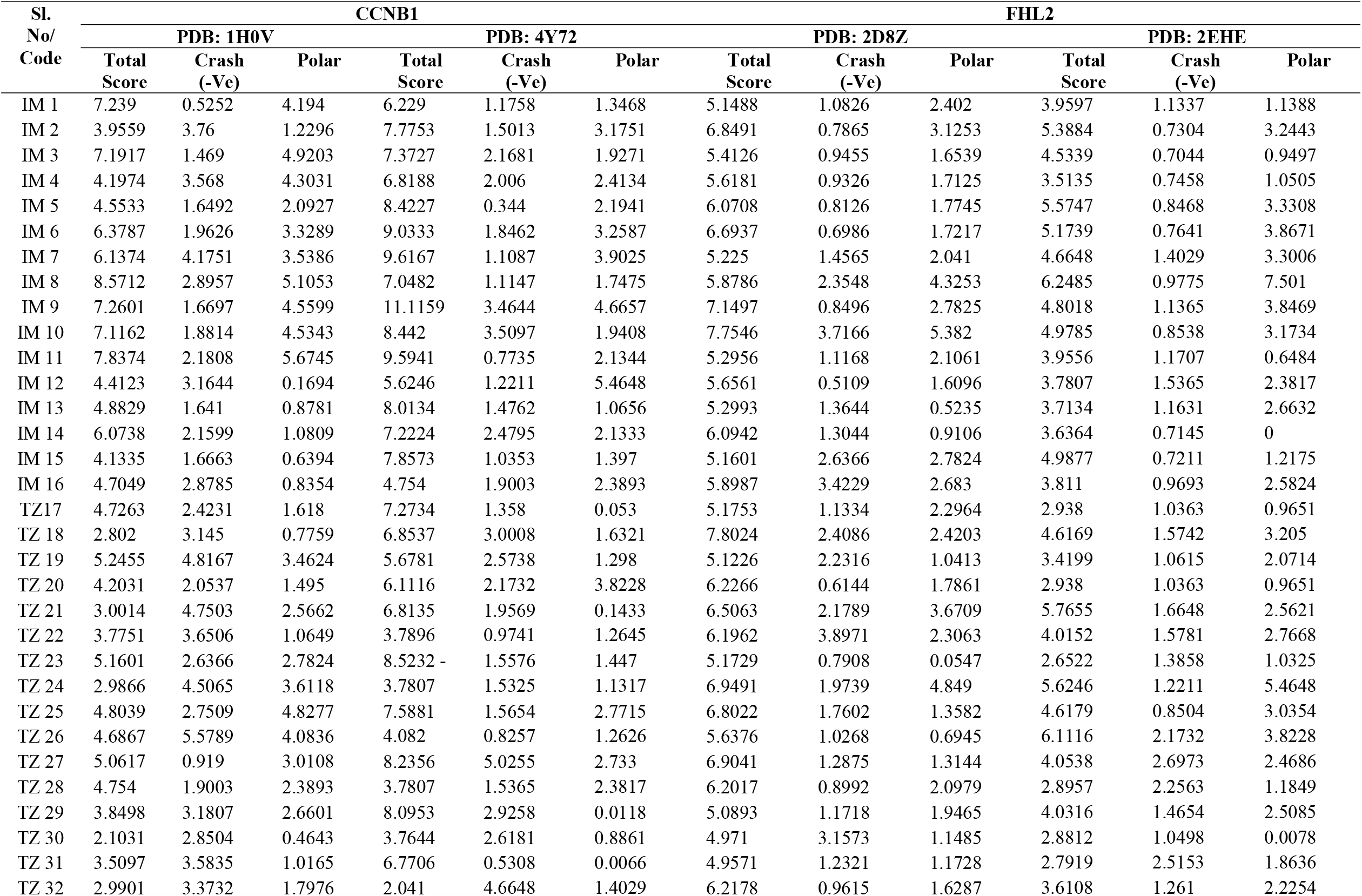
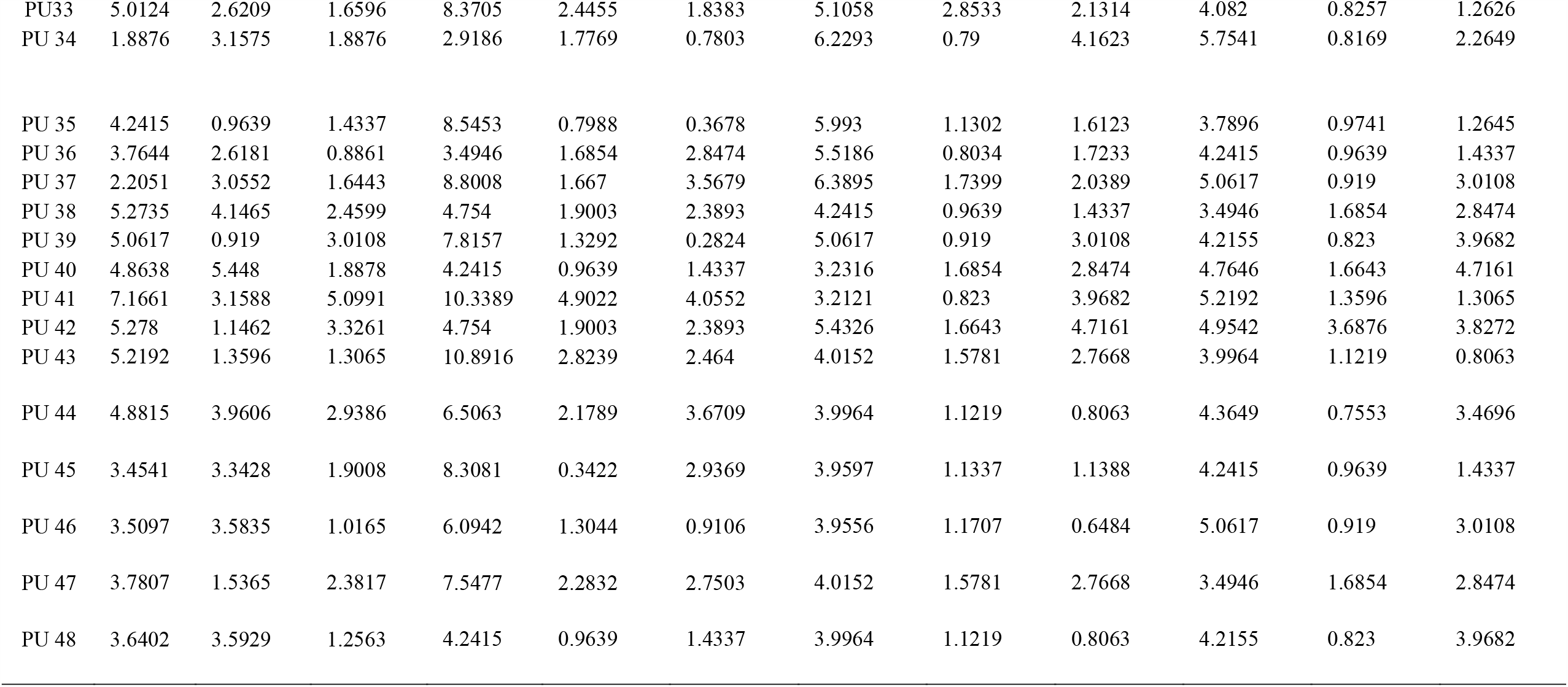
Docking results of designed molecules on CCNB1 and FHL2

| Sl.<br>No/<br>Code | CCNB1 |  |  |  |  |  | FHL2 |  |  |  |  |  |
| --- | --- | --- | --- | --- | --- | --- | --- | --- | --- | --- | --- | --- |
|  | PDB: 1H0V |  |  | PDB: 4Y72 |  |  | PDB: 2D8Z |  |  | PDB: 2EHE |  |  |
|  | Total<br>Score | Crash<br>(-Ve) | Polar | Total<br>Score | Crash<br>(-Ve) | Polar | Total<br>Score | Crash<br>(-Ve) | Polar | Total<br>Score | Crash<br>(-Ve) | Polar |
| IM 1 | 7.239 | 0.5252 | 4.194 | 6.229 | 1.1758 | 1.3468 | 5.1488 | 1.0826 | 2.402 | 3.9597 | 1.1337 | 1.1388 |
| IM 2 | 3.9559 | 3.76 | 1.2296 | 7.7753 | 1.5013 | 3.1751 | 6.8491 | 0.7865 | 3.1253 | 5.3884 | 0.7304 | 3.2443 |
| IM 3 | 7.1917 | 1.469 | 4.9203 | 7.3727 | 2.1681 | 1.9271 | 5.4126 | 0.9455 | 1.6539 | 4.5339 | 0.7044 | 0.9497 |
| IM 4 | 4.1974 | 3.568 | 4.3031 | 6.8188 | 2.006 | 2.4134 | 5.6181 | 0.9326 | 1.7125 | 3.5135 | 0.7458 | 1.0505 |
| IM 5 | 4.5533 | 1.6492 | 2.0927 | 8.4227 | 0.344 | 2.1941 | 6.0708 | 0.8126 | 1.7745 | 5.5747 | 0.8468 | 3.3308 |
| IM 6 | 6.3787 | 1.9626 | 3.3289 | 9.0333 | 1.8462 | 3.2587 | 6.6937 | 0.6986 | 1.7217 | 5.1739 | 0.7641 | 3.8671 |
| IM 7 | 6.1374 | 4.1751 | 3.5386 | 9.6167 | 1.1087 | 3.9025 | 5.225 | 1.4565 | 2.041 | 4.6648 | 1.4029 | 3.3006 |
| IM 8 | 8.5712 | 2.8957 | 5.1053 | 7.0482 | 1.1147 | 1.7475 | 5.8786 | 2.3548 | 4.3253 | 6.2485 | 0.9775 | 7.501 |
| IM 9 | 7.2601 | 1.6697 | 4.5599 | 11.1159 | 3.4644 | 4.6657 | 7.1497 | 0.8496 | 2.7825 | 4.8018 | 1.1365 | 3.8469 |
| IM 10 | 7.1162 | 1.8814 | 4.5343 | 8.442 | 3.5097 | 1.9408 | 7.7546 | 3.7166 | 5.382 | 4.9785 | 0.8538 | 3.1734 |
| IM 11 | 7.8374 | 2.1808 | 5.6745 | 9.5941 | 0.7735 | 2.1344 | 5.2956 | 1.1168 | 2.1061 | 3.9556 | 1.1707 | 0.6484 |
| IM 12 | 4.4123 | 3.1644 | 0.1694 | 5.6246 | 1.2211 | 5.4648 | 5.6561 | 0.5109 | 1.6096 | 3.7807 | 1.5365 | 2.3817 |
| IM 13 | 4.8829 | 1.641 | 0.8781 | 8.0134 | 1.4762 | 1.0656 | 5.2993 | 1.3644 | 0.5235 | 3.7134 | 1.1631 | 2.6632 |
| IM 14 | 6.0738 | 2.1599 | 1.0809 | 7.2224 | 2.4795 | 2.1333 | 6.0942 | 1.3044 | 0.9106 | 3.6364 | 0.7145 | 0 |
| IM 15 | 4.1335 | 1.6663 | 0.6394 | 7.8573 | 1.0353 | 1.397 | 5.1601 | 2.6366 | 2.7824 | 4.9877 | 0.7211 | 1.2175 |
| IM 16 | 4.7049 | 2.8785 | 0.8354 | 4.754 | 1.9003 | 2.3893 | 5.8987 | 3.4229 | 2.683 | 3.811 | 0.9693 | 2.5824 |
| TZ17 | 4.7263 | 2.4231 | 1.618 | 7.2734 | 1.358 | 0.053 | 5.1753 | 1.1334 | 2.2964 | 2.938 | 1.0363 | 0.9651 |
| TZ 18 | 2.802 | 3.145 | 0.7759 | 6.8537 | 3.0008 | 1.6321 | 7.8024 | 2.4086 | 2.4203 | 4.6169 | 1.5742 | 3.205 |
| TZ 19 | 5.2455 | 4.8167 | 3.4624 | 5.6781 | 2.5738 | 1.298 | 5.1226 | 2.2316 | 1.0413 | 3.4199 | 1.0615 | 2.0714 |
| TZ 20 | 4.2031 | 2.0537 | 1.495 | 6.1116 | 2.1732 | 3.8228 | 6.2266 | 0.6144 | 1.7861 | 2.938 | 1.0363 | 0.9651 |
| TZ 21 | 3.0014 | 4.7503 | 2.5662 | 6.8135 | 1.9569 | 0.1433 | 6.5063 | 2.1789 | 3.6709 | 5.7655 | 1.6648 | 2.5621 |
| TZ 22 | 3.7751 | 3.6506 | 1.0649 | 3.7896 | 0.9741 | 1.2645 | 6.1962 | 3.8971 | 2.3063 | 4.0152 | 1.5781 | 2.7668 |
| TZ 23 | 5.1601 | 2.6366 | 2.7824 | 8.5232 | 1.5576 | 1.447 | 5.1729 | 0.7908 | 0.0547 | 2.6522 | 1.3858 | 1.0325 |
| TZ 24 | 2.9866 | 4.5065 | 3.6118 | 3.7807 | 1.5325 | 1.1317 | 6.9491 | 1.9739 | 4.849 | 5.6246 | 1.2211 | 5.4648 |
| TZ 25 | 4.8039 | 2.7509 | 4.8277 | 7.5881 | 1.5654 | 2.7715 | 6.8022 | 1.7602 | 1.3582 | 4.6179 | 0.8504 | 3.0354 |
| TZ 26 | 4.6867 | 5.5789 | 4.0836 | 4.082 | 0.8257 | 1.2626 | 5.6376 | 1.0268 | 0.6945 | 6.1116 | 2.1732 | 3.8228 |
| TZ 27 | 5.0617 | 0.919 | 3.0108 | 8.2356 | 5.0255 | 2.733 | 6.9041 | 1.2875 | 1.3144 | 4.0538 | 2.6973 | 2.4686 |
| TZ 28 | 4.754 | 1.9003 | 2.3893 | 3.7807 | 1.5365 | 2.3817 | 6.2017 | 0.8992 | 2.0979 | 2.8957 | 2.2563 | 1.1849 |
| TZ 29 | 3.8498 | 3.1807 | 2.6601 | 8.0953 | 2.9258 | 0.0118 | 5.0893 | 1.1718 | 1.9465 | 4.0316 | 1.4654 | 2.5085 |
| TZ 30 | 2.1031 | 2.8504 | 0.4643 | 3.7644 | 2.6181 | 0.8861 | 4.971 | 3.1573 | 1.1485 | 2.8812 | 1.0498 | 0.0078 |
| TZ 31 | 3.5097 | 3.5835 | 1.0165 | 6.7706 | 0.5308 | 0.0066 | 4.9571 | 1.2321 | 1.1728 | 2.7919 | 2.5153 | 1.8636 |
| TZ 32 | 2.9901 | 3.3732 | 1.7976 | 2.041 | 4.6648 | 1.4029 | 6.2178 | 0.9615 | 1.6287 | 3.6108 | 1.261 | 2.2254 |
| PU33 | 5.0124 | 2.6209 | 1.6596 | 8.3705 | 2.4455 | 1.8383 | 5.1058 | 2.8533 | 2.1314 | 4.082 | 0.8257 | 1.2626 |
| PU 34 | 1.8876 | 3.1575 | 1.8876 | 2.9186 | 1.7769 | 0.7803 | 6.2293 | 0.79 | 4.1623 | 5.7541 | 0.8169 | 2.2649 |
| PU 35 | 4.2415 | 0.9639 | 1.4337 | 8.5453 | 0.7988 | 0.3678 | 5.993 | 1.1302 | 1.6123 | 3.7896 | 0.9741 | 1.2645 |
| PU 36 | 3.7644 | 2.6181 | 0.8861 | 3.4946 | 1.6854 | 2.8474 | 5.5186 | 0.8034 | 1.7233 | 4.2415 | 0.9639 | 1.4337 |
| PU 37 | 2.2051 | 3.0552 | 1.6443 | 8.8008 | 1.667 | 3.5679 | 6.3895 | 1.7399 | 2.0389 | 5.0617 | 0.919 | 3.0108 |
| PU 38 | 5.2735 | 4.1465 | 2.4599 | 4.754 | 1.9003 | 2.3893 | 4.2415 | 0.9639 | 1.4337 | 3.4946 | 1.6854 | 2.8474 |
| PU 39 | 5.0617 | 0.919 | 3.0108 | 7.8157 | 1.3292 | 0.2824 | 5.0617 | 0.919 | 3.0108 | 4.2155 | 0.823 | 3.9682 |
| PU 40 | 4.8638 | 5.448 | 1.8878 | 4.2415 | 0.9639 | 1.4337 | 3.2316 | 1.6854 | 2.8474 | 4.7646 | 1.6643 | 4.7161 |
| PU 41 | 7.1661 | 3.1588 | 5.0991 | 10.3389 | 4.9022 | 4.0552 | 3.2121 | 0.823 | 3.9682 | 5.2192 | 1.3596 | 1.3065 |
| PU 42 | 5.278 | 1.1462 | 3.3261 | 4.754 | 1.9003 | 2.3893 | 5.4326 | 1.6643 | 4.7161 | 4.9542 | 3.6876 | 3.8272 |
| PU 43 | 5.2192 | 1.3596 | 1.3065 | 10.8916 | 2.8239 | 2.464 | 4.0152 | 1.5781 | 2.7668 | 3.9964 | 1.1219 | 0.8063 |
| PU 44 | 4.8815 | 3.9606 | 2.9386 | 6.5063 | 2.1789 | 3.6709 | 3.9964 | 1.1219 | 0.8063 | 4.3649 | 0.7553 | 3.4696 |
| PU 45 | 3.4541 | 3.3428 | 1.9008 | 8.3081 | 0.3422 | 2.9369 | 3.9597 | 1.1337 | 1.1388 | 4.2415 | 0.9639 | 1.4337 |
| PU 46 | 3.5097 | 3.5835 | 1.0165 | 6.0942 | 1.3044 | 0.9106 | 3.9556 | 1.1707 | 0.6484 | 5.0617 | 0.919 | 3.0108 |
| PU 47 | 3.7807 | 1.5365 | 2.3817 | 7.5477 | 2.2832 | 2.7503 | 4.0152 | 1.5781 | 2.7668 | 3.4946 | 1.6854 | 2.8474 |
| PU 48 | 3.6402 | 3.5929 | 1.2563 | 4.2415 | 0.9639 | 1.4337 | 3.9964 | 1.1219 | 0.8063 | 4.2155 | 0.823 | 3.9682 |

**Fig. 13.**
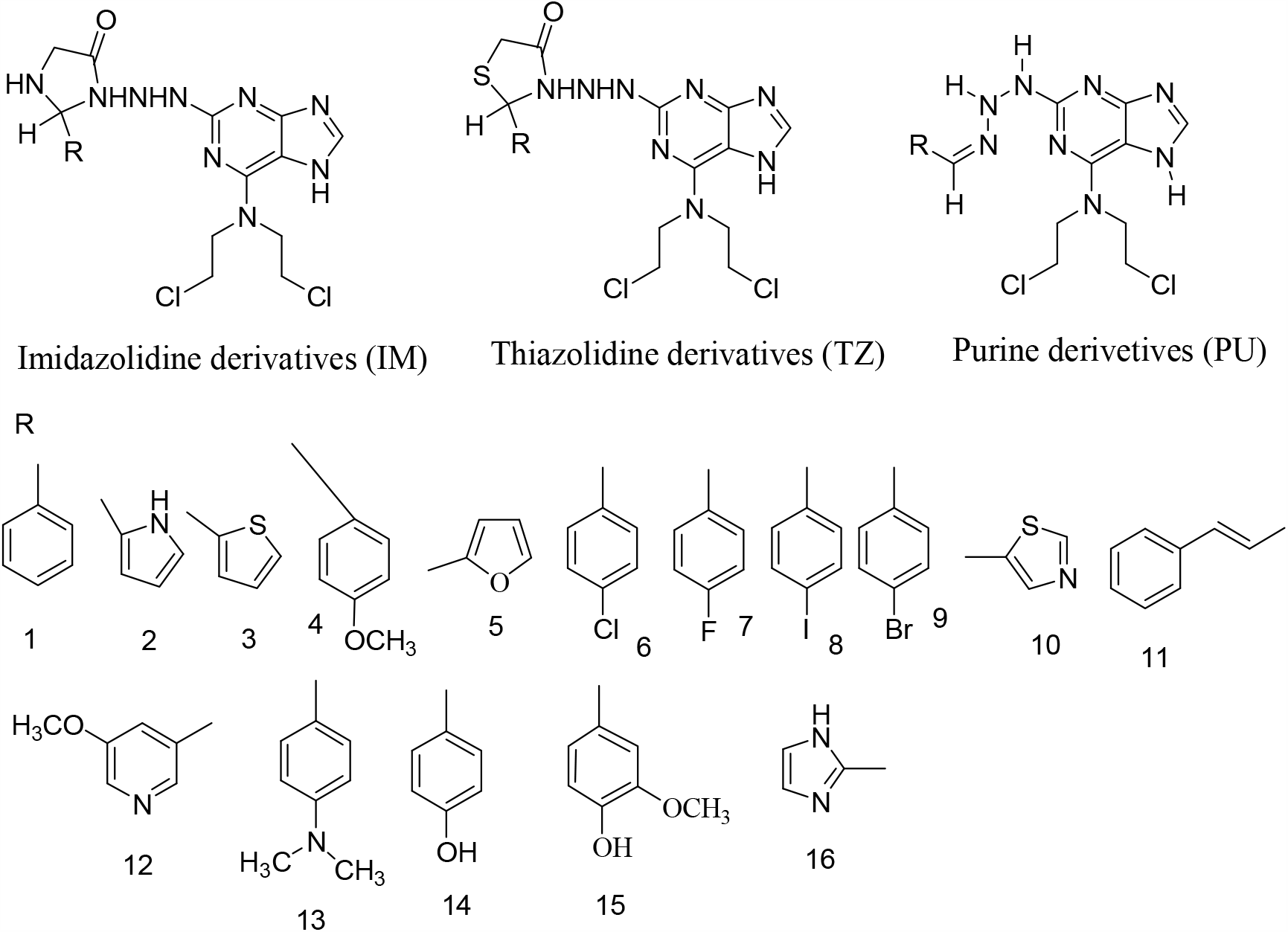
Structures of Designed Molecules

**Fig. 14.**
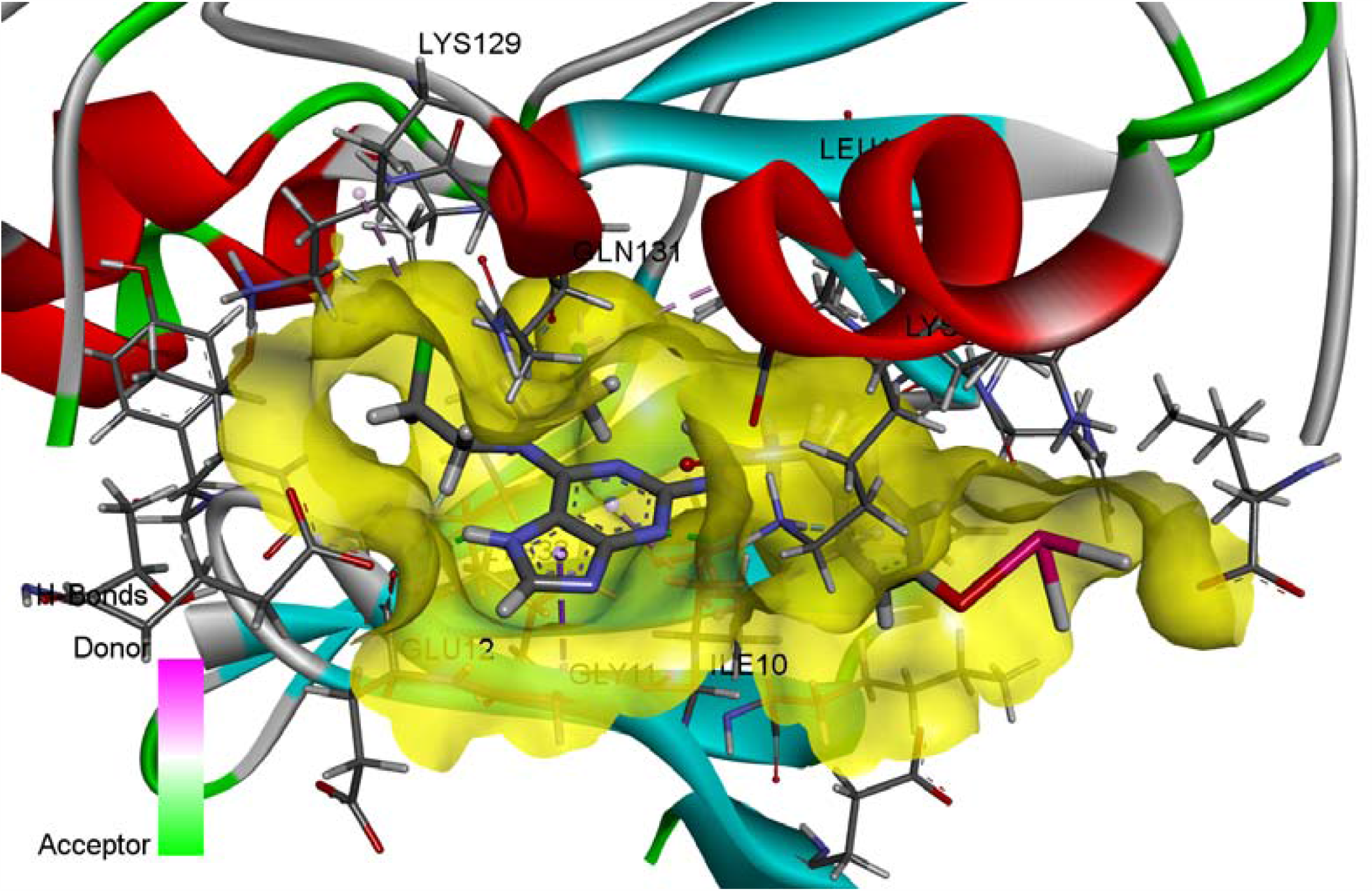
3D Binding of Molecule IM 8 with 1H0V

**Fig. 15.**
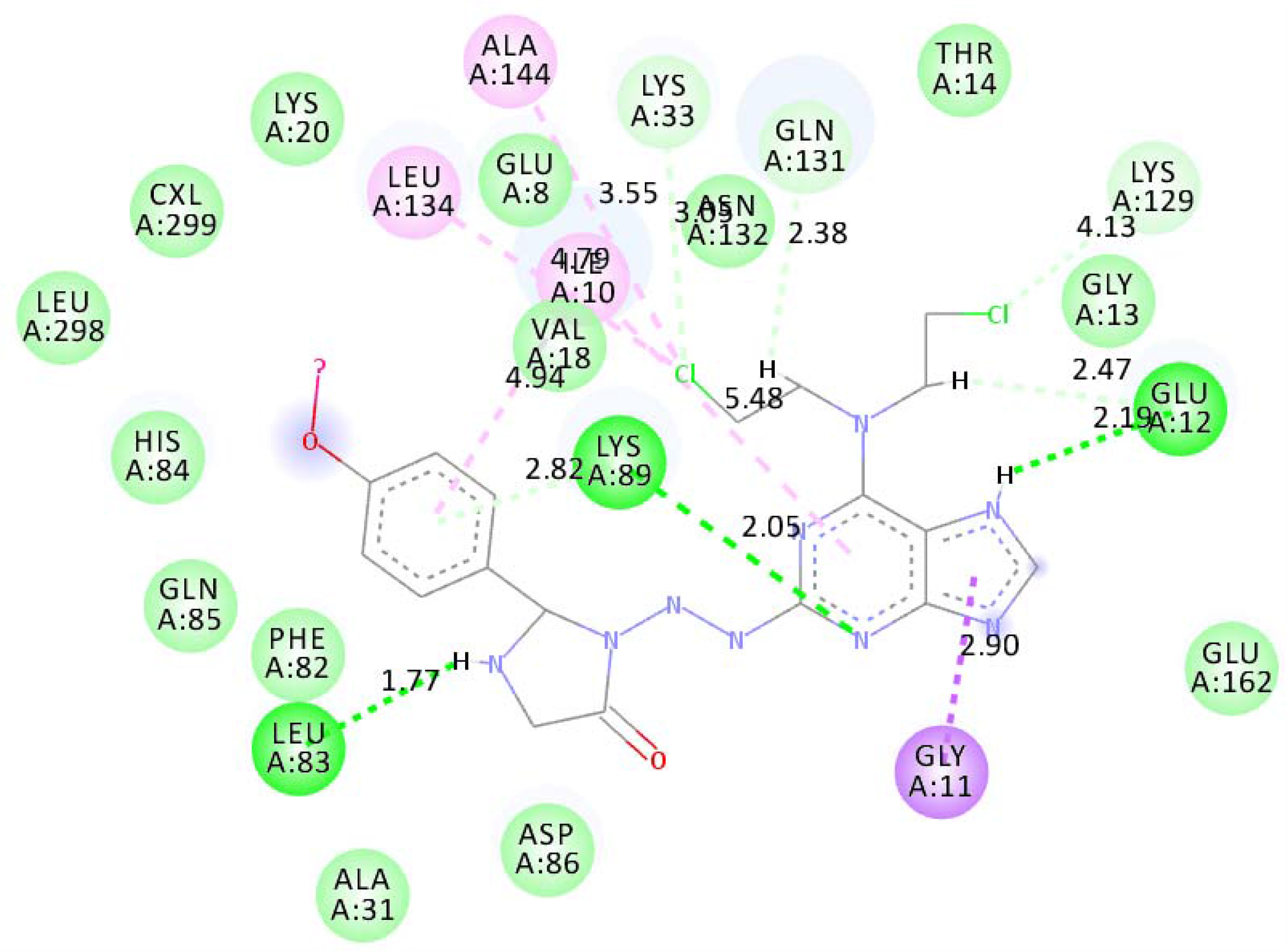
2D Binding of Molecule IM 8 with 1H0V

## Discussion

Due to the high heterogeneity of PDAC, PDAC was still a disease with high rates of pervasiveness and fatality. With surgery as the main, the other treatments including radiotherapy, chemotherapy, targeted therapy, and gene therapy as a additive to the finite treatment measures of PDAC, the 5-year survival rate was still less than 8% [35]. Therefore, the early diagnosis and effective treatment of PDAC is crucially required, which may be achieved via the identification of the DEGs between PDAC and normal control, and by considerate the underlying molecular mechanism. Microarray and high throughput sequencing analysis can screen a massive number of genes in the human genome for farther functional analysis, and can be extensively used to screen biomarkers for early diagnosis and unique therapeutic targets. Therefore, they may help the diagnosis of PDAC in the early stages and the advancement of targeted treatment, thus developing prognosis.

The current investigation systematically applied integrated bioinformatics methods to identify novel biomarkers that serve roles in the advancement PDAC. The data extracted from the GEO dataset contained 31 pairs of lung cancer and normal samples. A total of 232 up regulated and 231 down regulated genes in PDAC, when compared with normal control samples, were identified using bioinformatics analysis, indicating that the incidence and advancement of PDAC. The results of the DEGs may provide potential biomarkers for the diagnosis of PDAC. DAP (death associated protein) [36], KRT8 [37], IGFBP2 [38], KRT19 [39], CD44 [40], AHNAK (AHNAK nucleoprotein) [41] and BTG1 [42] are a noticeable factors in the PDAC progression. Wang et al [43] reported that KIF2C induces proliferation, migration, and invasion in gastric cancer patients through the MAPK signaling pathway, but this gene might be associated with development of PDAC. DBN1 expression was significantly increased in breast cancer [44], but this gene might be liable for development of PDAC. MAP1B was reported to lung cancer progression [45], but this gene may be key for advancement of PDAC. BNIP3L down regulation was required to develop breast and ovarian cancer [46], but this down regulation of gene might be involved in progression of PDAC. Yen et al [47] reported that ITGA4 was expressed in oral cancer, but this gene might be novel biomarker for PDAC. Tomsic et al. [48] showed that mutation in SRRM2 was associated with progression of thyroid carcinoma, but alteration in this gene might be key factor for advancement of PDAC. Recent studies have shown that down regulation of IL7R is associated with progression of esophageal squamous cell carcinoma [49], but this gene might be involved in pathogenesis of PDAC. Lee et al [50] found that reduced expression of the HLA-DRA is key factor for development of colorectal cancer, but this gene might be linked with advancement of PDAC. Liu et al. [51] reported that the absence of SESN3 linked with development of hepatocellular carcinoma, but this gene might be associated with progression of PDAC.

Then, GO and REACTOME pathway analyses were used to investigate the interactions of these DEGs. Increasing evidence shows that LAPTM4B [52], CEACAM6 [53], SERPINE2 [54] and VNN1 [55], SPHK1 [56], HRG (histidine rich glycoprotein) [57], VEGFC (vascular endothelial growth factor C) [58], ANXA3 [59], APOA2 [60], LCN2 [61], TIMP1 [62], CD63 [63], CD151 [64], MAL2 [65], ARNTL2 [66], PKD2 [67], E2F1 [68], MMP1 [69], CCR7 [70], NOTCH2 [71], BTLA (B and T lymphocyte associated) [72], TFRC (transferrin receptor) [73], CD4 [74], ATM (ATM serine/threonine kinase) [75], LEF1 [76], CSF1R [77], CTSB (cathepsin B) [78], DUSP2 [79] and NR4A1 [80] are closely associated with progression of PDAC. PTGER3 [81] and MAGI2 [82] are linked with angiogenesis, chemoresistance, cell proliferation and migration in ovary cancer, but these genes might be liable for growth PDAC. Hoagland et al [83] demonstrated that HP (haptoglobin) expression was responsible for progression of lung cancer, but this gene might be involved in PDAC progression. FGA (fibrinogen alpha chain) was demonstrated to be a lung cancer susceptibility gene through activation of integrin–AKT signaling pathway [84], but this gene might be liable for progression of PDAC. Repetto et al [85] investigated the importance of FGB (fibrinogen beta chain) in the pathogenesis of gastric carcinoma, but this gene might be responsible for progression of PDAC. PLA2G4A [86], FGG (fibrinogen gamma chain) [87] and TYMS (thymidylatesynthetase) [88] have been demonstrated to be up regulated in cancer, but these genes might be liable for progression of PDAC. RAB32 [89], SEPTIN4 [90], TPM2 [91], ACOT7 [92], PRTFDC1 [93], CABLES1 [94], HLA-DMB [95], PTPRC (protein tyrosine phosphatase receptor type C) [96], CD5 [97], CD6 [97], MS4A1 [98], CD22 [99], CD27 [100], MRC2 [101], CLEC2D [102], EEF1A1 [103] and APOB (apolipoprotein B) [104] played a predominant role in the cancer progression, but these genes might be associated with development of PDAC. Jung et al [105] found that SMPD1 stimulates the drug resistance in colorectal cancer, but this gene might be linked with development of PDAC. Liu et al. [106], Yang et al [107], Song et al [108], Seachrist et al [109], Zhu et al [110], Wu et al [111], Wang et al [112], Yi et al [113], Lan et al [114] and Appert-Collin et al [115] revealed that PADI4, MAOB (monoamine oxidase B), TRPC6, BCL11A, CXCR5, TCF7, POU2F2, SLC4A1, STK17B and LRP1 were associated with cancer cell invasion, but these genes might be liable for progression of PDAC. Kairouz et al [116], Diez-Bello et al [117], Xue et al [118], Abo-Elfadl et al [119], Li et al [120] and Zhao et al [121] reported that GRB14, TRPC6, ZFPM2, TNFRSF13B, ADAM19 and PIK3IP1enhance the cancer cell proliferation, but this gene might be involved in advancement of PDAC. Leite et al [122], Feng et al [123], Wang et al [124], Zhong et al [125], Yokoyama-Mashima et al [126], Guo et al [127], Lawson et al [128] and Wang et al [129] demonstrated that low levels of HLA-DPA1, FGL2, CBLB (Cbl proto-oncogene B), NCKAP1L, DYRK2, OGT (O-linked N-acetylglucosamine (GlcNAc) transferase), CAMK1D and RNF213 are linked with progression of cancer, but these genes might be essential for progression of PDAC. Polymorphic genes such as RORA (RAR related orphan receptor A) [130], IGF2R [131] and ZBTB20 [132] are contribute to progression of cancer, but these genes might be crucial for advancement of PDAC. Hope et al. [133] identified that the VCAN (versican) was central role in immune cell infiltration in cancer, but this gene may be associated with immune cell infiltration in PDAC.

Construction of PPI network and modules of DEGs may be helpful for understanding the relationship of developmental PDAC. Bai et al [134] reported that CCNB1 plays a positive role in proliferation of cancer cells, but this gene might be involved in development of PDAC. FHL2 [135] and RPL10 [136] are associated with progression of PDAC. Further investigation is required in order to clarify the underlying biological mechanisms of novel biomarkers HLA-DPA1, TUBB1, RPL13A, RPL27A and RPL23A on PDAC.

The miRNA-DEG regulatory network and TF-DEG regulatory network were constructed to explore the molecular mechanism of PDAC. The EZH2 [137], KMT2D [138], TXNIP (thioredoxin interacting protein) [139], TP63 [140], SOX2 [141], MYC [142] and KLF4 [143] genes are associated with PDAC. TPM1 [144] and hsa-mir-1202 [145] have been associated with cancer risk, but these genes and miRNAs might be responsible for progression of PDAC. Hsa-mir-4461, hsa-mir-3928-3p and hsa-mir-2682-5p might be considered as novel biomarkers for progression of PDAC.

In conclusion, we aim to identify DEGs by bioinformatics analysis to find the potential biomarkers which may be involved in the advancement of PDAC. The investigation contributes a set of useful DEGs for future studies into molecular mechanisms and biomarkers of PDAC. And the application of data mining and integration is accessible for prediction of PDAC advancement. Nevertheless, further molecular biological analyses are recommended to certify the function of the DEGs in PDAC.

## Ethical approval

This article does not contain any studies with human participants or animals performed by any of the authors.

## Informed consent

No informed consent because this study does not contain human or animals participants.

## Availability of data and materials

The datasets supporting the conclusions of this article are available in the GEO (Gene Expression Omnibus) (https://www.ncbi.nlm.nih.gov/geo/) repository. [(GSE133684) (https://www.ncbi.nlm.nih.gov/geo/query/acc.cgi?acc=GSE133684]

## Consent for publication

Not applicable.

## Acknowledgement

I thank Shenglin Huang, Fudan University, Shanghai, China, very much, the author who deposited their microarray dataset, GSE133684, into the public GEO database.

## Author Contributions

Basavaraj Vastrad-Writing original draft, and review and editing

Anandkumar Tengli **-** Writing original draft and investigation

Chanabasayya Vastrad - Software and investigation

